# Immune Cell Deformability in Depressive Disorders: Longitudinal Associations Between Depression, Glucocorticoids and Cell Deformability

**DOI:** 10.1101/2022.09.23.22280275

**Authors:** Andreas Walther, Martin Kräter, Clemens Kirschbaum, Wei Gao, Magdalena Wekenborg, Marlene Penz, Nicole Rothe, Jochen Guck, Lucas Daniel Wittwer, Julian Eder

## Abstract

**Background:** Cell deformability of all major blood cell types is increased in depressive disorders (DD). Furthermore, impaired glucocorticoid secretion is causally related to DD. Nevertheless, there are no longitudinal studies examining changes in glucocorticoid output and depressive symptoms regarding cell deformability in DD.

**Aim:** To investigate, whether changes in depressive symptoms or hair glucocorticoids predict cell deformability in DD.

**Methods:** In 136 individuals, depressive symptoms (PHQ-9) and hair glucocorticoids (cortisol and cortisone) were measured at timepoint one (T1), while one year later (T2) depressive symptoms and hair glucocorticoids were remeasured and additionally cell deformability of peripheral blood cells was assessed and DD status was determined by clinical interview.

**Results:** Depression severity at T1 predicted higher cell deformability in monocytes and lymphocytes over the entire sample. Subjects with continuously high depressive symptoms at T1 and T2 showed elevated monocyte deformability as compared to subjects with low depressive symptoms. Depression severity at T1 of subjects with a lifetime persistent depressive disorder (PDD) was associated with elevated monocyte, neutrophil, and granulo-monocyte deformability. Depression severity at T1 of subjects with a 12-month PDD was positively associated with monocyte deformability. Furthermore, increases in glucocorticoid concentrations from T1 to T2 tended to be associated with higher immune cell deformability, while strongest associations emerged for the increase in cortisone with elevated neutrophil and granulo-monocyte deformability in the 12-month PDD group.

**Conclusion:** Continuously elevated depressive symptomatology as well as an increase in glucocorticoid levels over one year are associated with higher immune cell deformability, particularly in PDD. These findings suggest, that persistent depressive symptomatology associated with increased glucocorticoid secretion may lead to increased immune cell deformability thereby compromising immune cell function and likely contributing to the perpetuation of PDD.

## Introduction

Of all disorders, depressive disorders (DD) cause the greatest disease burden and disability worldwide (World Health Organisation (WHO), 2017). Major depressive disorder (MDD) is characterized by a period of at least two weeks of depressive mood or anhedonia in combination with other symptoms such as appetite changes, insomnia/hypersomnia, increased fatigue, or feelings of worthlessness (American Psychiatric Association, 2013a). Persistent depressive disorder (PDD; formerly dysthymia) is characterized on the basis of depressed mood over a two-year period in combination with symptoms similar to MDD, although PDD is often perceived as less severe in terms of intensity (American Psychiatric Association, 2013a). Nevertheless, both conditions cause severe functional impairment and there are major difficulties in reducing the high prevalence rates (Hasin et al., 2018; Vos et al., 2017) and improving DD treatment (Munder et al., 2019; Munkholm et al., 2019), with 33% of patients not achieving remission despite multiple treatment attempts (Guidi et al., 2011; Rush et al., 2006). Often, the poor therapeutic efficacy is attributed to the still insufficiently understood pathophysiology of DD (Biselli et al., 2019; Fiacco et al., 2019; Otte et al., 2016; Rothe et al., 2020).

Research increasingly recognizes that key pathophysiological processes occur directly at the cellular level and that DD can impair peripheral blood cell function via disturbed glucocorticoid secretion by the hypothalamus-pituitary-adrenal (HPA) axis or via inflammation (Lopez-Vilchez et al., 2016; Moylan et al., 2013; Pariante, 2017; Rodrigues et al., 2014; Walther et al., 2018; Wolkowitz et al., 2010). Blood cells fulfill multiple functions from primary immune response, over metabolite transport to overall blood flow. Real-time deformability cytometry (RT-DC) representing a new method to investigate the physical properties of blood cells such as cell mechanical features or cell size provides unprecedented insights into overall blood cell function (Otto et al., 2015a; Rosenbluth et al., 2008a). It has been highlighted that the assessment of the mechanical state of blood cells, measured by cell deformability, is appropriate to detect and classify human diseases conditions such as spherocytosis, malaria, or COVID-19 infections (Kubánková et al., 2021; Toepfner et al., 2018a) and is predictive for immune cell activation (Bashant et al., 2020, 2019).

Only recently, using RT-DC, we were able to show that in DD and especially in PDD immune cell deformability is increased as compared to healthy controls (Walther et al., 2022b). Although all major blood cells tended to be more deformable, lymphocytes, monocytes and neutrophils were particularly affected indicating immune cell mechanical changes to occur in DD. This is potentially related to disturbed HPA axis function or dysfunctional immune response. In addition, optical traps (Ekpenyong et al., 2012), atomic force microscopy (Lam et al., 2008; Rosenbluth et al., 2008b), and micropipette aspiration (Ravetto et al., 2014) have been used in proof-of-concept studies to reveal mechanical changes in immune cells under pathological conditions. Nevertheless, due to the general dominance of erythrocytes in blood and the consequent much increased statistical power to detect associations, previous studies investigated only the relationship between erythrocyte deformability and mental disorders (Jasenovec et al., 2019; Saha et al., 2019). The study by Saha et al. (2019) reported patients with chronic fatigue syndrome / myalgic encephalomyelitis to exhibit lower erythrocyte deformability than healthy controls. Further, Jasenovec et al. (2019) examined children within the autism spectrum identifying those children with more severe symptoms to show impaired erythrocyte deformability. It should be noted here, however, that the current gold standard for measuring cell deformability, namely RT-DC (Kräter et al., 2021; Nawaz et al., 2020; Otto et al., 2015a; Urbanska et al., 2020; Wu et al., 2018), was only used in our landmark study comparing individuals with DD and healthy controls (Walther et al., 2022b).

In contrast, there is a large body of literature examining the relationship between impaired glucocorticoid secretion by the HPA axis and DD (Rothe et al., 2020). Indeed, it seemed to be widely accepted that MDD is associated with increased HPA activity and increased basal cortisol concentrations in the circulation (Otte et al., 2016; Stetler and Miller, 2011). However, due to the inconclusive findings on cortisol stress reactivity in MDD, this is no longer definitive (Miller and Kirschbaum, 2019; Rothe et al., 2020; Zorn et al., 2017). Furthermore, investigating new tissue sources such as hair cortisol showing mixed associations with regard to DD highlight on the one hand the relevance of the investigation of chronic glucocorticoid output over longer time periods (Gerritsen et al., 2019; Rothe et al., 2021, 2020; Stalder et al., 2017; Steudte-schmiedgen et al., 2017; Walther et al., 2022a), but on the other hand, it emerges as increasingly important to investigate the effects of impaired chronic glucocorticoid secretion on cell function. Studies increasingly show that the extracellular microenvironment is critical to modify cellular physiology, which ultimately determines cell functionality (Kim et al., 2019). So, understanding the interactions between immune cells and DD-related alterations in glucocorticoid secretion appears to be increasingly important for understanding differences in immune cell deformability in health and disease.

In addition to increased cortisol levels and the often-identified chronic low-grade inflammation in individuals suffering from DD (Pariante, 2017), Lynall et al. (2019) further reported elevated numbers of immune cells in depressed individuals compared to controls. It is known that increased levels of glucocorticoids and catecholamines lead to increased immune cell count, as cells demarginate from the vessel walls. Interestingly, these observations were associated with cellular softening (Fay et al., 2016). It is suggested that the underlying mechanism leading to increased immune cell counts in depressed individuals is rooted in the effect of elevated glucocorticoid levels remodeling the actin cytoskeleton of blood cells and thereby softening leukocytes and enabling them to demarginate from the vessel wall (Fay et al., 2016). Not only starts the actin cytoskeleton to be remodeled with continuously elevated levels of glucocorticoids, but also are lipid metabolism and composition crucially affected resulting in increased softening and bending of blood cells (Demirkan et al., 2013; Knowles et al., 2017; Liu et al., 2016; Ravetto et al., 2014; Walther et al., 2018). This cascade of processes ultimately impairs blood cell function and may be an underlying cause of symptoms of fatigue and exhaustion in DD. The disturbed immune cell function in connection with subsequent dysregulated cytokine release could trigger sickness behavior with anhedonia and fatigue (Dantzer, 2009).

Therefore, we hypothesize that continuously high levels of depressive symptoms are associated with increased immune cell deformability. We further hypothesize that high and especially continuously high glucocorticoid levels as well as an increase in glucocorticoid secretion over one year is associated with elevated immune cell deformability in individuals with DD.

## Method

### Study design

This observational study used a longitudinal design with a baseline measurement and a one-year follow-up measurement. Cross-sectional results of the study were reported elsewhere (Walther et al., 2022b). Baseline examination of participants took place between October and December 2018, while follow-up examinations were conducted one year later. The study, entitled Mood-related Morpho-rheological Changes in Peripheral Blood Cells (Mood-Morph), used a participant pool of a large-scale prospective cohort study to identify eligible participants and obtain pre-assessed baseline data (Penz et al., 2018). Eligible participants were asked via e-mail whether they were willing to participate in an add-on study to the prospective longitudinal study.

For the study, participants with elevated depressive symptoms, as measured by the Patient Health Questionnaire (PHQ-9), and gender- and age-matched healthy controls were identified from the participant pool. These participants provided for the baseline measurement (T1) sociodemographic information, depressive symptomatology and further health-related data. Also, collected hair samples for glucocorticoid quantification were obtained for the study at T1. At the follow-up time point (T2), the psychometric assessment and hair sampling was repeated complemented with a clinical diagnostic interview to diagnose depression status and blood samples were obtained to measure cell deformability. The study was approved by the local ethics committee of the Dresden University of Technology (EK182042019) and all participants gave written informed consent to participate in the study.

### Participants

For study inclusion, participants’ age had to fall between 18 and 68 years, and interested study participants were excluded if they had any kind of blood disorder that could affect blood cell deformability. Potential participants who were contacted to participate in the Mood-Morph study, were informed in the study invitation and informational email that certain diseases, such as spherocytosis and blood-related diseases in general, precluded participation (Toepfner et al., 2018a). Potential participants with a cold or other acute infections no longer ago than two weeks, were not included to exclude acutely infectious participants. Acute medical conditions such as blood diseases, cancer, severe cardiovascular diseases, or pregnancy led to exclusion, while in the population very prevalent conditions such as hypertension or hypothyroidism did not lead to exclusion. Differing conditions were recorded and used as covariate in the analyses as well as the specific drug categories (e.g., for hypertension—antihypertensive drugs).

In terms of depression levels measured with the PHQ-9, only subjects with a T1 PHQ-9 score >10 (high risk of depression) as well as age- and sex-matched subjects with a PHQ-9 score <5 (low risk of depression) were included. The aim of this procedure was to obtain a sample of participants with high and low depressive symptoms at baseline and to be able to identify change in depressive symptoms over one year. At T2, participants underwent a clinical diagnostic interview leading to exclusion of participants from the DD groups in case participants did not fulfill diagnostic criteria, had a positive mania screening, or suffered from another major psychological disorder. Exclusion from the healthy control group occurred if participants exhibited a major psychological disorder or psychopharmacological medication intake.

### Procedures

Participants provided sociodemographic and health-related information (e.g., gender, age, medical conditions, medication) as well as depressive symptom severity and information on their general health online via the study-homepage platform. By using a personalized code, participants accessed a private study space, signed first consent forms regarding data privacy, clinical data collection and saving, and hair/blood sampling and completed subsequently their study questionnaires. For depression severity the PHQ-9 and for general health the Short Form Health Questionnaire (SF-12) was completed (Kroenke et al., 2001; Ware Jr et al., 1996). While the SF-12 measures general physical and psychological health, the PHQ-9 is a nine-item self-report questionnaire for the measurement of depression severity rating the frequency of DSM-5 diagnostic criteria for MDD (e.g., feeling down, depressed, or hopeless) during the past two weeks on a four-point Likert scale (0 = not at all, 3 = nearly every day). Scores of 10 or greater were consistently shown to identify with high sensitivity and specificity cases of MDD (He et al., 2020; Levis et al., 2020). Since the scale developers suggested that PHQ-9 scores of 5, 10, 15, and 20 represent mild, moderate, moderately severe, and severe depression (Kroenke et al., 2001), individuals with scores below 5 were considered to have minimal depressive disorder risk, whereas individuals with scores of 15 or higher were considered to have high depressive disorder risk.

To obtain capillary blood and scalp hair samples, participants arrived at the Dresden University of Technology at the Department of Biopsychology, where trained personnel performed blood and hair sampling. At T2, after the bio-sampling took place, the Composite International Diagnostic Interview (DIA-X-5/CIDI) was conducted by trained interviewers (Hoyer et al., 2020). The DIA-X-5 is a standardized clinical interview for the assessment of mental disorders. The DIA-X-5 assesses lifetime symptoms and past 12 months symptoms. To rule out bipolar disorders, a mania-screening questionnaire, consisting of the initial questions of the DIA-X-5 mania section, was conducted to detect any lifetime (hypo-) mania-symptoms. Lifetime and 12-month diagnoses were subsequently generated for PDD and MDD according to DSM-5 criteria (American Psychiatric Association, 2013b). After finishing the clinical interview, subjects received 15 € compensation for expenses. Duration of the examination at T2 was approximately one hour.

### Real-time deformability cytometry (RT-DC)

A 20 µl capillary blood sample was extracted from the fingertip of participants using a safety lancet (Safety-Lancet Normal 21, Sarstedt, Nümbrecht, Germany) and harvested in a capillary (Minivette POCT, 20 µl, Sarstedt, Nümbrecht, Germany). The blood was immediately resuspended in 380 µl RT-DC measurement buffer containing 0.6% methylcellulose in phosphate buffered saline (CellCarrierB, Zellmechanik Dresden, Germany) in a microcentrifuge tube. Blood-samples were then transferred to the Department of Cellular Machines at the Biotechnology Center of the TU Dresden, where the samples were measured using an RT-DC device. Maintained at room temperature samples were measured within three hours since sampling according to a protocol published elsewhere (Toepfner et al., 2018b).

In brief, blood was flushed through a microfluidic channel constriction 20 µm x 20 µm in cross section (Flic20, Zellmechanik Dresden, Germany) by applying a constant flow rate. An image of every measured blood cell was taken by a high-speed camera and cell deformability and cell size were calculated (Otto et al., 2015b). RT-DC measurements were controlled by the acquisition software Shape-In2 (Zellmechanik Dresden, Germany). The different blood cell types were classified by utilizing artificial intelligence-based image classification as published elsewhere (Kräter et al., 2021) and mean values for cell deformability and cell size of every donor and blood cell type were extracted.

### Hair sampling and liquid chromatography and mass-spectrometry (LC-MS)

Three hair strands (each containing at least 20 mg of hair) were cut as close as possible to the scalp from the posterior vertex for quantification of hair cortisol and cortisone. Hair sample processing and analysis were performed as described in our published protocols for glucocorticoids (Gao et al., 2016, 2013). Preprocessing of the samples and subsequent biochemical analysis were performed by Dresden LabService GmbH (Tatzberg 47, 01307, Dresden, Germany). Samples were cut into 3 cm segments from the scalp site representing the integrated hormone concentration over the last three months (e.g., average hair growth rate of 1 cm per month; (Wennig, 2000)) and weighed into 7.5 mg of whole, non-pulverized hair and then washed with 2.5 ml of isopropanol according to the protocol of Gao et al. (Gao et al., 2013). Biochemical analysis was performed by liquid chromatography coupled with tandem mass spectrometry (LC-MS/MS) as described in detail elsewhere (Gao et al., 2016). The intra- and inter-assay coefficients of variation for cortisol and cortisone are less than 8.8% and 8.5%, respectively. The lower limit of detection for cortisol and cortisone is 0.09 pg/mg.

### Statistical analysis

Statistical analyses were run in R v4.2.1 (R Core Team, 2017). Data was checked for normality using Shapiro-Wilk-tests and for variance homogeneity using *F* tests. In case of violation of normal distribution, non-parametric tests were conducted. Differences in confounding variables (sex, age, BMI, psychopharmaceutical intake, medication category) and in PHQ-9, cortisol, cortisone and WBC count were tested between subjects suffering from depression and healthy controls using two-tailed independent *t*-tests (Student’s or Welch two sample *t*-test), Mann-Whitney *U* tests, univariate analysis of variance (ANOVA) and *χ2*-tests in dependence of normal distribution and scale of measurement. Pearson and Kendall correlation coefficients (Croux and Dehon, 2010) were tested to check for possible (non-)linearity relationships. Partial correlations for T1 and T2 data were conducted to control for gender, age, BMI and psychopharmaceutical treatment, while for change scores (T2-T1) a baseline T1 correction (e.g., PHQ-9 T1 or cortisol (or cortisone) T1) was additionally applied. All *p*-values were adjusted using Holm-Bonferroni correction for multiple comparisons (Holm, 1979). Cortisol and cortisone data from both measurements were log-transformed due to their non-normal distribution and tested two-tailed. Correlation analyses were performed over the entire sample size and in each subgroup with differing depressive disorders.

## Results

### Sample description

In total, 136 (*n_female_* = 100; *n_male_* = 36) individuals, who completed the PHQ-9 and provided hair samples for glucocorticoid quantification at T1 and T2 were used for subsequent analyses investigating the longitudinal association between depressive symptoms, glucocorticoid secretion and immune cell deformability. Moreover, the selected participants (*N* = 136) were assigned to different diagnostic groups in dependence of their DIA-X diagnoses. Of initially 139 participants, due to incomplete PHQ-9 data of three individuals, following group size emerged from the diagnostic classification for the 136 participants: Lifetime PDD (*n* = 30), 12-month PDD (*n* = 15), lifetime MDD (*n* = 40), 12-month MDD (12-month MDD; *n* = 11) and healthy controls (HC; *n* = 61). Participants suffering from double depression were not treated as a separate group due to small sample size and lack of statistical power. A detailed representation of the sample flow is provided in Figure 1. Sample characteristics of the entire sample and the different diagnostic groups is presented in Table 1 and Supplementary Table 1. With respect to hair cortisol and cortisone analyses, sample size of the initially entire sample was used (*N* = 139). However, as hair segments could not be obtained from all subjects due to lack of sufficient hair of some participants, sample size varies within each group. Regarding cortisol analyses, one outlier was additionally excluded due to a hair cortisol concentration above 100 pg/mg. For cortisol at T1 and T2 n = 116 observations were available at each time point for analysis, while for cortisone at T1 and T2 n = 118 observations were available at each time point.

**Figure 1.**
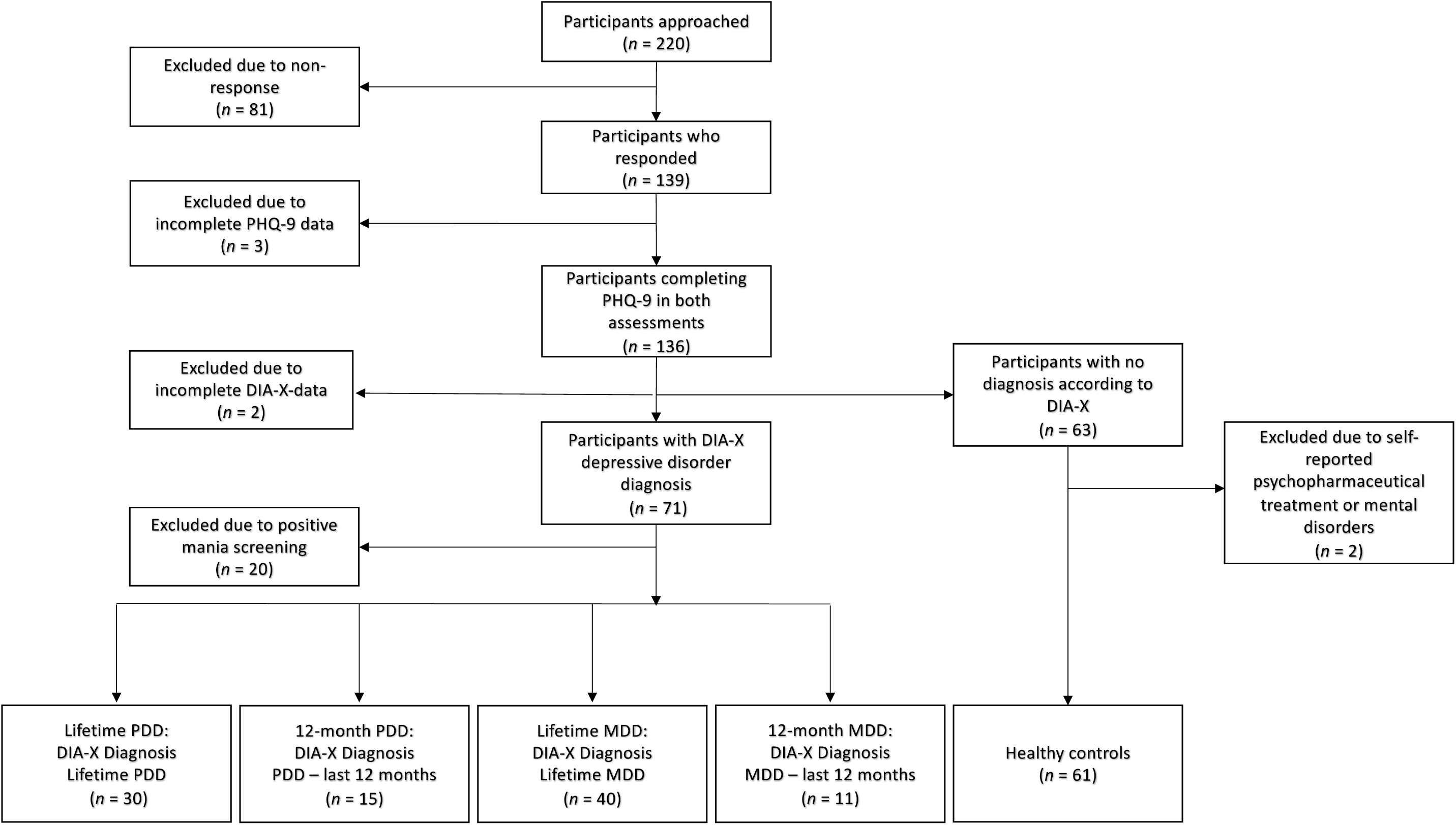
Sample Flow Leading to Relevant Case-Control Groups

**Table 1.** Sample characteristics according to the entire sample

| Sample characteristics | Entire sample (n = 136) | Range (min – max) |
| --- | --- | --- |
| Age<br>*M (SD)* | 46.72 (11.28) | 20 – 65 |
| Sex<br>*females (%)* | 100 (73.53%) | – |
| Body mass index<br>*M (SD)* | 25.51 (4.88) | 17.57 – 40.09 |
| Psychopharmaceutical<br>intake<br>*frequency* | 23 (16.91%) | – |
| Medication<br>*frequency* | 74 (54.41%) | – |
| PHQ-9 T1<br>*M (SD)* | 8.74 (6.98) | 0 – 25 |
| PHQ-9 T2<br>*M (SD)* | 7.65 (6.03) | 0 – 25 |
| Hair cortisol T1 (pg/mg)<br>*M (SD)* | 18.35 (8.90) | 1.75 – 37.00 |
| Hair cortisol T2 (pg/mg)<br>*M (SD)* | 10.60 (10.15) | 1.00 – 71.55 |
| Hair cortisone T1 (pg/mg)<br>*M (SD)* | 27.38 (18.35) | 4.71 – 90.39 |
| Hair cortisone T2 (pg/mg)<br>*M (SD)* | 32.28 (22.86) | 6.51 – 93.22 |

*Note.* T1 = time point 1, T2 = time point 2, PHQ-9 = Patient Health Questionnaire 9, n = number of participants, M = mean, SD = standard deviation, number of hair cortisol observations at T1 and T2 = 116, number of hair cortisone observations T1 and T2 = 118.

### Control of Confounders

Participants in subgroups with differing depressive disorders showed no significant differences regarding age, sex and BMI compared to healthy controls. Expected significant group differences were observed in PHQ-9 (T1 and T2, T2-T1) scores and psychopharmaceutical treatment within all groups except PHQ-9 symptom change score (T2-T1) for the 12-month PDD group and controls. Exact group differences of the diagnostic groups are shown in Supplementary Table 1. Subgroups with differing depressive disorders and healthy controls did not show a significant difference in glucocorticoid concentrations (T1, T2, T2-T1; see Supplementary Tables 2 and 3 and Supplementary Figures 1–6). No significant correlations between depressive symptoms (PHQ-9 T1 and PHQ-9 T2) and mean cell area size over the entire sample were found in any cell type (see Supplementary Table 4 and 5). No significant differences in immune cell count were found between participants with a DD diagnosis and healthy controls (see Supplementary Table 6). Furthermore, cell deformability did not differ with respect to the six different medication groups (see Supplementary Text 1).

### Association Between Depression Severity and Cell Deformability

Pearson, Kendall and Partial correlation coefficients between PHQ-9 (T1) scores and cell deformability for each cell type are depicted in Table 2. Below, only significant Pearson correlations are described. Significant correlations between PHQ-T1 and monocytes were found for the entire sample (r = .22, p = .0051) and the lifetime PDD group (r = .33, p = .0049) both corrected for confounder variables (entire sample: r_p_ = .20, p = .01; lifetime PDD: r_p_ = .32, p = .0085) and applied to Holm-Bonferroni correction (entire sample: p = .0245, p_p_ = .0425; lifetime PDD: p = .0245, p_p_ = .0425). Furthermore, within the lifetime PDD group, significant correlations between PHQ-9 scores (T1) and neutrophil deformability (r = .30, p = .0097, r_p_ = .33, p = .0072) as well as granulo-monocytes (r = .32, p = .0057; r_p_ = .34, p = .0057) emerged, indicating higher immune cell deformability in subjects, who suffered from PDD (see Figure 2). Pearson, Kendall and partial correlations remained significant after Holm-Bonferroni correction (p = .0485, p_p_ = .036). No further correlations did survive correction for multiple testing. Nevertheless, with respect to the entire sample a significant correlation between PHQ-9 (T1) and lymphocyte deformability was observed (r = .16, p = .0326; r_p_ = .17, p = .0270). Within the 12-month PDD group, significant Pearson and Kendall correlations between PHQ-9 scores (T1) and monocyte deformability were found (r = .35, p = .0306), but not when confounders were taken into account. Within the 12-month PDD group, a significant positive partial correlation with respect to erythrocyte deformability was found (r = .36, p = .0353). No stable significant correlations emerged for the lifetime MDD and 12-month MDD group. Significant results indicate a positive correlation of PHQ-9 scores at T1 with immune cell deformability in the lifetime PDD group and partially over the entire sample. Correlations between PHQ-9 scores (T2) and cell deformability for each cell type was performed over the entire sample revealing significant correlations for monocyte cell deformability (r = .17, p = .0254; r_p_ = 16, p = .036; see Supplementary Table 7). Because of the limited variance between T1 and T2 of PHQ-9 scores, we did not focus on the association of depressive symptoms difference scores and cell deformability. Nevertheless, an overview of the correlations can be seen in the Supplementary Table 8.

**Figure 2.**
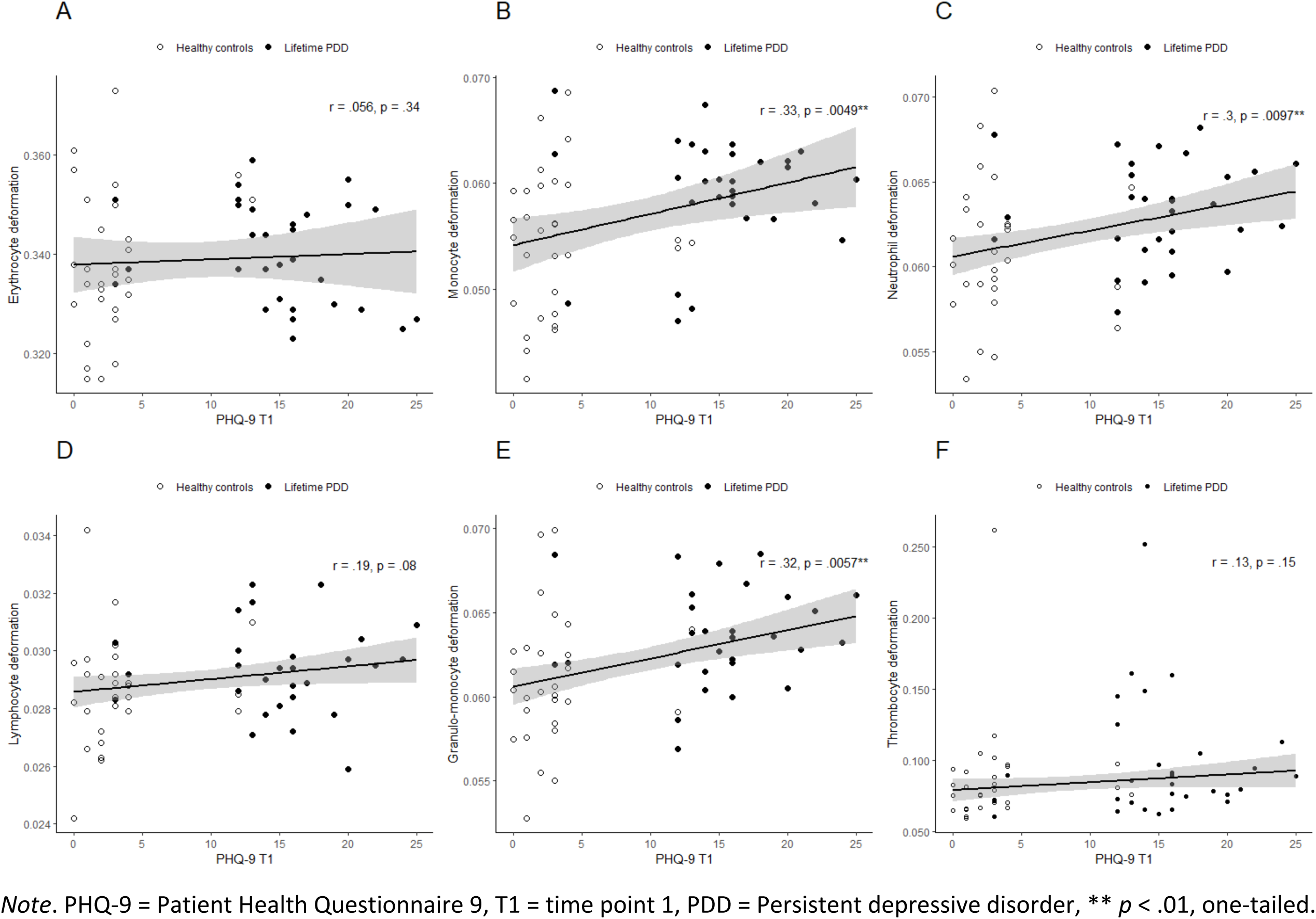
Associations of Depressive Symptoms Measured by PHQ-9 (T1) and Cell Deformability According to Lifetime PDD group

**Table 2.**
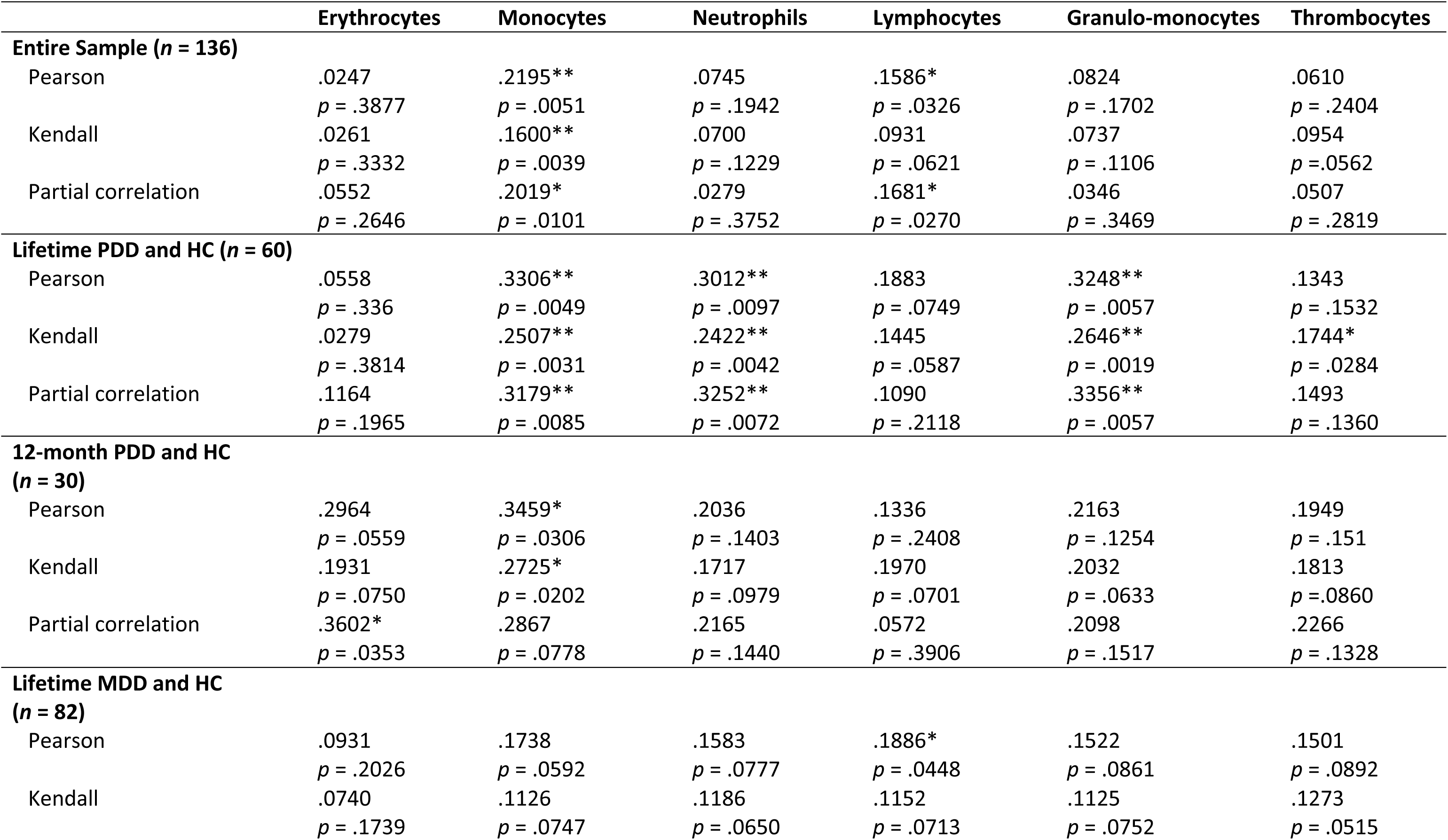
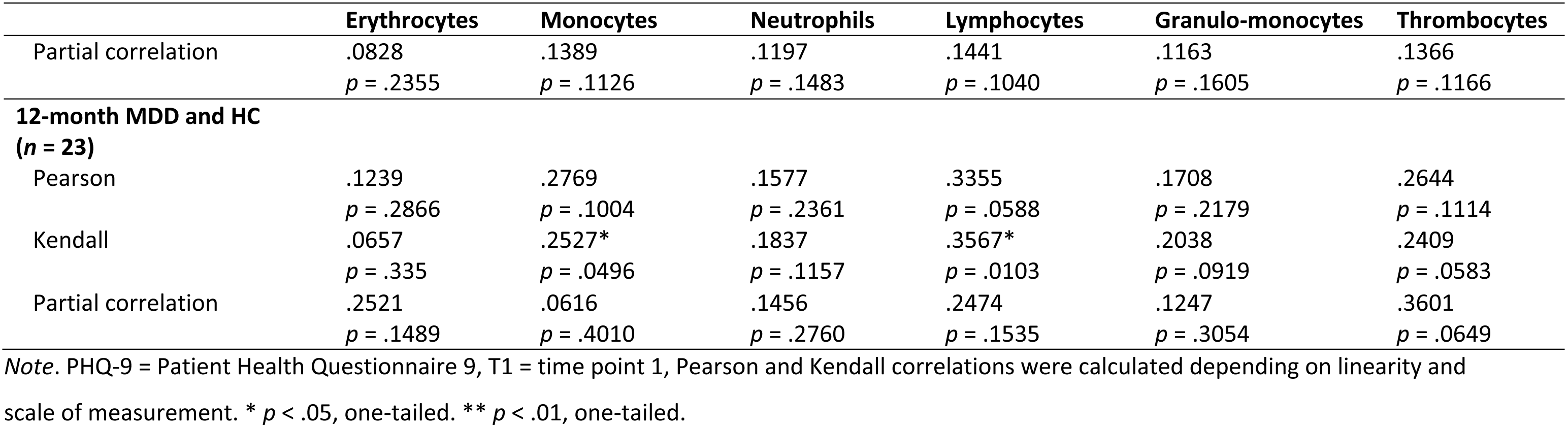
Associations Between Depressive Symptoms Measured by PHQ-9 (T1) and Cell Deformability

### Association Between Cortisol and Cortisone (T1 and T2) and Cell Deformability

No significant association between cortisol at T1 and immune cell deformability of any cell type was revealed over the entire sample or in any subgroup (see Supplementary Table 9). Similarly, between cortisol level T2 and immune cell deformability no significant correlation emerged (see Supplementary Table 10). Also, no significant association between cortisone level T1 and immune cell deformability emerged over the entire sample or in any subgroup (see Supplementary Table 11). A single significant Pearson correlation between cortisone T2 and lymphocyte deformability was observed in the 12-month PDD group (*r* = .38, *p* = .0419) (*n* = 29), which did not survive correction for multiple testing. No further significant correlations were observed between cortisone T2 and immune cell deformability for any cell type in any subgroup (see Supplementary Table 12).

Overall, no stable significant correlations between cortisol, cortisone (T1 and T2) and immune cell deformability were observed for the entire sample nor in any subgroup.

### Association Between *Cortisol and Cortisone Change (T2-T1)* and Cell Deformability

To examine the association between cortisol or cortisone change from T1 to T2 and cell deformability independent of group affiliation, Pearson and Kendall correlations were computed. Significant correlations between cortisol and cortisone difference (T2-T1), respectively, and neutrophil deformability as well as granulo-monocyte deformability emerged for the lifetime PDD group (see Figure 3) and the 12-month PDD group. The results indicate that in those groups, subjects with an increase in cortisol or cortisone over the two assessments exhibit higher neutrophil or granulo-monocyte deformability at T2. Significant partial correlations remained when controlling for gender, age, BMI, psychopharmaceutical treatment and baseline cortisol or cortisone levels (T1) in the 12-month PDD group. However, only the association between cortisone difference (T2-T1) and neutrophil and granulo-monocyte deformability survived Holm-Bonferroni correction in the 12-month PDD group (neutrophil deformability: *p* = .0455, *p_p_* = .0062; granulo-monocyte deformability: *p* = .019, *p_p_* = .0034). No stable significant correlations for lifetime MDD and 12-month MDD were observed. An overview of correlations is presented in Table 3 and Table 4.

**Figure 3.**
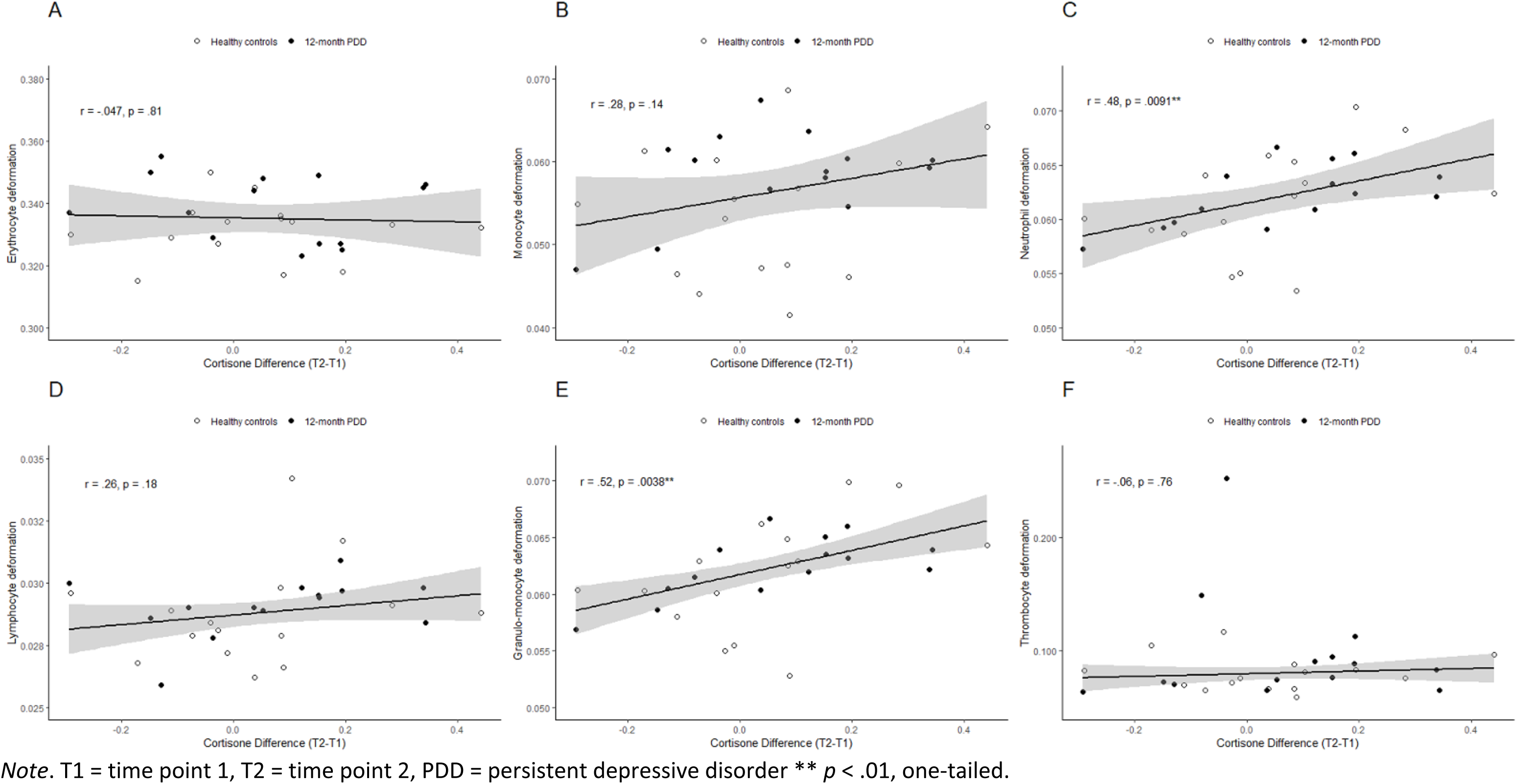
Associations of Cortisone Difference (T2-T1) and Cell Deformability According to 12-month PDD group

**Table 3.** Associations Between Cortisol Difference (T2-T1) and Cell Deformability

|  | Erythrocytes | Monocytes | Neutrophils | Lymphocytes | Granulo-Monocytes | Thrombocytes |
| --- | --- | --- | --- | --- | --- | --- |
| **Entire Sample (n = 116)** |  |  |  |  |  |  |
| Pearson | .0580<br>*p* = .5366 | .0560<br>*p* = .5501 | -.0194<br>*p* = .8365 | -.1343<br>*p* = .1505 | -.0233<br>*p* = .8038 | -.1676<sup>†</sup><br>*p* = .0722 |
| Kendall | -.0080<br>*p* = .8993 | .0296<br>*p* = .6383 | .0128<br>*p* = .8393 | -.0646<br>*p* = .3080 | .0107<br>*p* = .8654 | -.1223<sup>†</sup><br>*p* = .0518 |
| Partial correlation | .0618<br>*p* = .5197 | 0.0436<br>*p* = 0.6494 | -.0107<br>*p* = 0.9109 | -0.1422<br>*p* = .1366 | -.0202<br>*p* = .8334 | -.1458<br>*p* = .1269 |
| **Lifetime PDD and HC (n = 50)** |  |  |  |  |  |  |
| Pearson | -.1052<br>*p* = .4671 | .0760<br>*p* = .6000 | .2921*<br>*p* = .0396 | .0575<br>*p* = .6914 | .2898*<br>*p* = .0412 | .0190<br>*p* = .8961 |
| Kendall | -.0669<br>*p* = .4974 | .0418<br>*p* = .6695 | .2298*<br>*p* = .0191 | .0774<br>*p* = .4313 | .2434*<br>*p* = .0129 | .0613<br>*p* = .5304 |
| Partial correlation | -.1507<br>*p* = .3230 | .0459<br>*p* = .7646 | .2729<br>*p* = .0698 | .0935<br>*p* = .5411 | .2645<br>*p* = .0791 | .0699<br>*p* = .6484 |
| **12-month PDD and HC (n = 29)** |  |  |  |  |  |  |
| Pearson | -.3028<br>*p* = .1104 | .1032<br>*p* = .5944 | .3546<sup>†</sup><br>*p* = .0591 | .2500<br>*p* = .1908 | .3862*<br>*p* = .0385 | .0345<br>*p* = .8589 |
| Kendall | -.2644*<br>*p* = .0463 | .0865<br>*p* = .5112 | .2343<sup>†</sup><br>*p* = .0747 | .1814<br>*p* = .1703 | .3189*<br>*p* = .0155 | .1603<br>*p* = .2227 |
| Partial correlation | -.3665<sup>†</sup><br>*p* = .0781 | -.0467<br>*p* = .8283 | .3990<sup>†</sup><br>*p* = .0534 | .2270<br>*p* = .2861 | .4162*<br>*p* = .0431 | .0391<br>*p* = .8561 |
| **Lifetime MDD and HC (n = 71)** |  |  |  |  |  |  |
| Pearson | .1027<br>*p* = .3940 | .0020<br>*p* = .9865 | -.0360<br>*p* = .7654 | -.1531<br>*p* = .2025 | -.0455<br>*p* = .7063 | -.0269<br>*p* = .8241 |
| Kendall | .0297<br>*p* = .7169 | -.0056<br>*p* = .9446 | .0073<br>*p* = .9288 | -.0292<br>*p* = .7206 | .0109<br>*p* = .8934 | .0085<br>*p* = .917 |
| Partial correlation | .1076<br>*p* = .3897 | .0129<br>*p* = .9179 | -.0056<br>*p* = .9647 | -.1451<br>*p* = .2450 | -.0193<br>*p* = .8780 | .06422<br>*p* = .6084 |
| **12-month MDD and HC**<br>**(*n* = 24)** |  |  |  |  |  |  |
| Pearson | .0342<br>*p* = .8738 | .3849<sup>†</sup><br>*p* = .0633 | .2335<br>*p* = .2721 | .1074<br>*p* = .6176 | .2711<br>*p* = .2001 | .1407<br>*p* = .5121 |
| Kendall | -.0511<br>*p* = .7280 | .2650<sup>†</sup><br>*p* = .0701 | .2214<br>*p* = .1301 | .1095<br>*p* = .4562 | .2826<sup>†</sup><br>*p* = .0555 | .1851<br>*p* = .2057 |
| Partial correlation | .1708<br>*p* = .4845 | .2599<br>*p* = .2826 | .2529<br>*p* = .2963 | .0030<br>*p* = .9902 | .2627<br>*p* = .2772 | .0469<br>*p* = .8489 |

Note. T1 = time point 1, T2 = time point 2, Pearson and Kendall tests were calculated depending on linearity and scale of measurement. \* *p* < .05, two-tailed. \*\* *p* < .01, two-tailed. <sup>†</sup> *p* < .1, two-tailed (trend).

**Table 4.** Associations Between Cortisone Difference (T2-T1) and Cell Deformability

|  | Erythrocytes | Monocytes | Neutrophils | Lymphocytes | Granulo-Monocytes | Thrombocytes |
| --- | --- | --- | --- | --- | --- | --- |
| **Entire Sample (n = 118)** |  |  |  |  |  |  |
| Pearson | - | .0713 | .0252 | -.0366 | .0138 | -.1706<sup>†</sup> |
|  | *p* = .9997 | *p* = .4426 | *p* = .7862 | *p* = .6942 | *p* = .8824 | *p* = .0647 |
| Kendall | -.0357 | .0202 | .0366 | -.0161 | .0230 | -.1330* |
|  | *p* = .5701 | *p* = .7464 | *p* = .4477 | *p* = .7980 | *p* = .7132 | *p* = .0329 |
| Partial correlation | .0116 | .0509 | .0415 | -.0389 | .0248 | -.1335 |
|  | *p* = .9033 | *p* = .5925 | *p* = .6622 | *p* = .6824 | *p* = .7945 | *p* = .1587 |
| **Lifetime PDD and HC (n = 51)** |  |  |  |  |  |  |
| Pearson | -.0248 | .1276 | .2679<sup>†</sup> | -.0089 | .2655<sup>†</sup> | -.1298 |
|  | *p* = .8627 | *p* = .3724 | *p* = .0574 | *p* = .9504 | *p* = .0597 | *p* = .3641 |
| Kendall | -.0525 | .0583 | .2176* | .0016 | .2299* | .0008 |
|  | *p* = .5913 | *p* = .5476 | *p* = .0249 | *p* = .987 | *p* = .0197 | *p* = .9935 |
| Partial correlation | .1253 | .0709 | .2641<sup>†</sup> | .0228 | .2560<sup>†</sup> | -.0928 |
|  | *p* = .4068 | *p* = .6396 | *p* = .0761 | *p* = .8805 | *p* = .0859 | *p* = .5398 |
| **12-month PDD and HC (n = 29)** |  |  |  |  |  |  |
| Pearson | -.0470 | .2810 | .4760** | .2581 | .5204** | -.0603 |
|  | *p* = .8071 | *p* = .1398 | *p* = .0091 | *p* = .1764 | *p* = .0038 | *p* = .756 |
| Kendall | -.1397 | .1609 | .03625** | .2162 | .4277** | .1159 |
|  | *p* = .2925 | *p* = .2224 | *p* = .0058 | *p* = .1022 | *p* = .0012 | *p* = .3779 |
| Partial correlation | -.0839 | .1554 | .6196** | .2594 | .6434*** | .0038 |
|  | *p* = .6968 | *p* = .4685 | *p* = .0012 | *p* = .2209 | *p* = .0007 | *p* = .9861 |
| **Lifetime MDD and HC (n = 73)** |  |  |  |  |  |  |
| Pearson | .0429 | -.0035 | -.0044 | -.0834 | -.0196 | -.1090 |
|  | *p* = .7188 | *p* = .9763 | *p* = .9706 | *p* = .4831 | *p* = .8689 | *p* = .3585 |
| Kendall | .0168 | -.0107 | .0210 | -.0491 | .0092 | -.0339 |
|  | *p* = .8339 | *p* = .8939 | *p* = .7933 | *p* = .5419 | *p* = .9090 | *p* = .6717 |
| Partial correlation | .0572<br>$p = .6433$ | -.0146<br>$p = .9060$ | .0109<br>$p = .9295$ | -.0901<br>$p = .4649$ | -.0087<br>$p = .9440$ | -.0598<br>$p = .6281$ |
| **12-month MDD and HC**<br>**($n = 24$)** |  |  |  |  |  |  |
| Pearson | .0898<br>$p = .6766$ | .3297<br>$p = .1156$ | .2977<br>$p = .1576$ | -.0543<br>$p = .8011$ | .3357<br>$p = .1087$ | .1141<br>$p = .5956$ |
| Kendall | .0730<br>$p = .6193$ | .2069<br>$p = .1573$ | .2505<sup>†</sup><br>$p = .0869$ | -.0146<br>$p = .9209$ | .2971*<br>$p = .0436$ | .0762<br>$p = .6023$ |
| Partial correlation | .1156<br>$p = .6375$ | .2482<br>$p = .3056$ | .3466<br>$p = .1460$ | -.0913<br>$p = .7100$ | .3686<br>$p = .1205$ | .1076<br>$p = .6611$ |

Note. Pearson and Kendall tests were calculated depending on linearity and scale of measurement. All correlations were calculated two-tailed. \* $p < .05$ , two-tailed. \*\* $p < .01$ , two-tailed, \*\*\* $p < .001$ , two-tailed, <sup>†</sup> $p < .1$ , two-tailed (trend).

### Cell Deformability Comparisons According to high and minimal DD risk Groups

Comparing the groups with high vs. minimal DD risk at T1 (high: PHQ-9 ≥ 15; minimal: PHQ-9 < 5), Welch two sample *t*-tests revealed a significant difference in monocyte deformability at T2 between the high risk group (*M* = .0585, *SD* = .0041) and the minimal risk group (*M* = .0549, *SD* = .0069; *t*(75.238) = −3.0642, *p* = .0015) indicating a moderate effect size (*d* = .62; see Figure 4). A significant difference in lymphocyte deformability at T2 between the high-risk group (*M* = .0294, *SD* = .0017) and minimal risk group (*M* = .0287, *SD* = .0018) was revealed by a Mann-Whitney *U* test (*U* = 677.5, *p* = .0337), indicating a small effect size (*r* = −.19). Furthermore, subjects in the consistent high risk group exhibiting at both time points PHQ-9 scores equal or above 15 (*M* = .0589, *SD* = .0035) showed higher monocyte deformability compared to the consistent minimal risk group exhibiting at both time points PHQ-9 scores lower than 5 (*M* = .0550, *SD* = .0067; *t*(37.138) = −2.9601, *p* = .0027) revealing a moderate effect size (*d* = .74; Figure 5). No further significant differences were found for any other cell type (see Supplementary Table 13).

**Figure 4.**
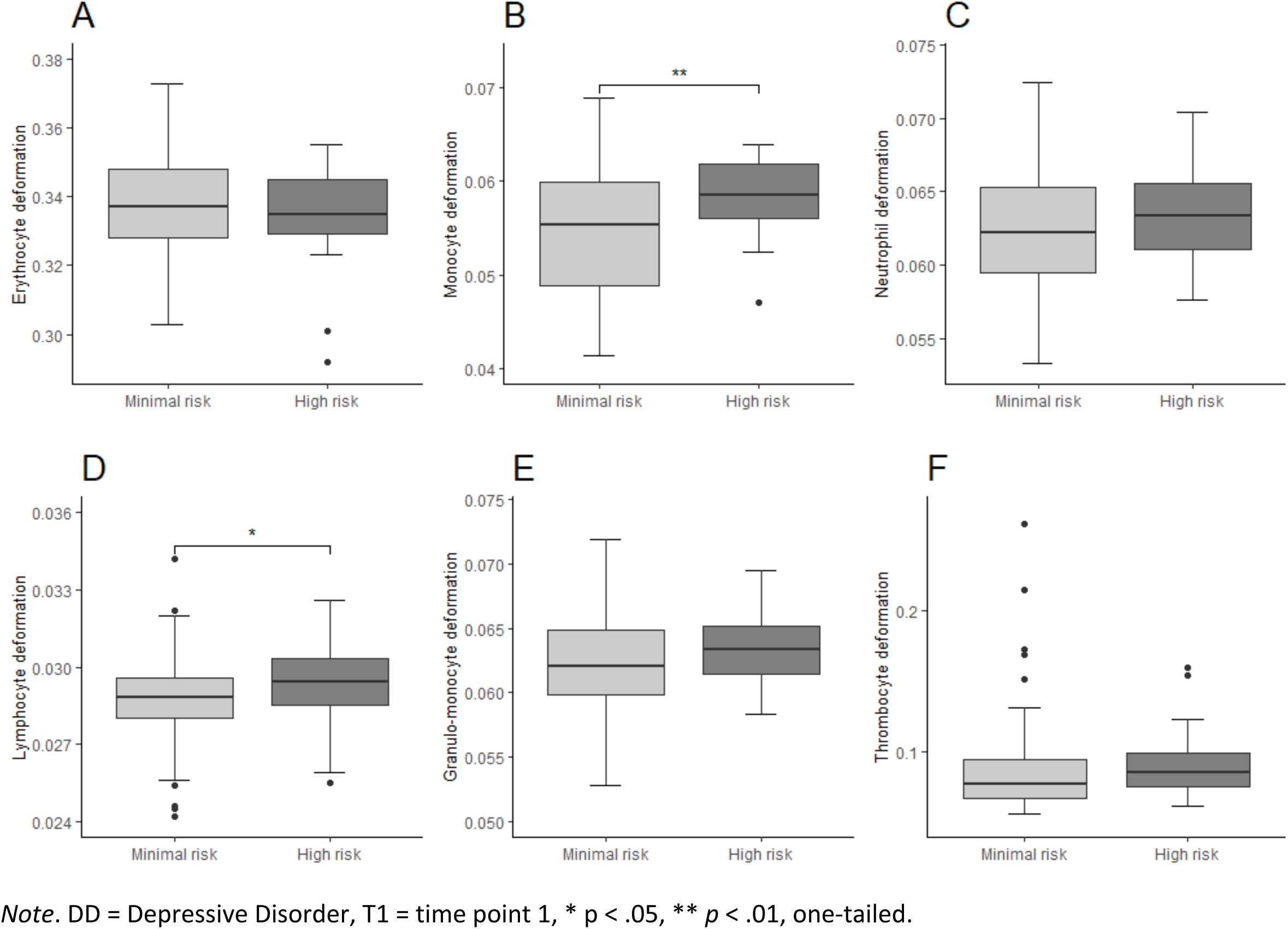
Combined Boxplots for the High DD Risk Group and the Minimal DD Risk Group (T1) Regarding Blood Cell Deformability

**Figure 5.**
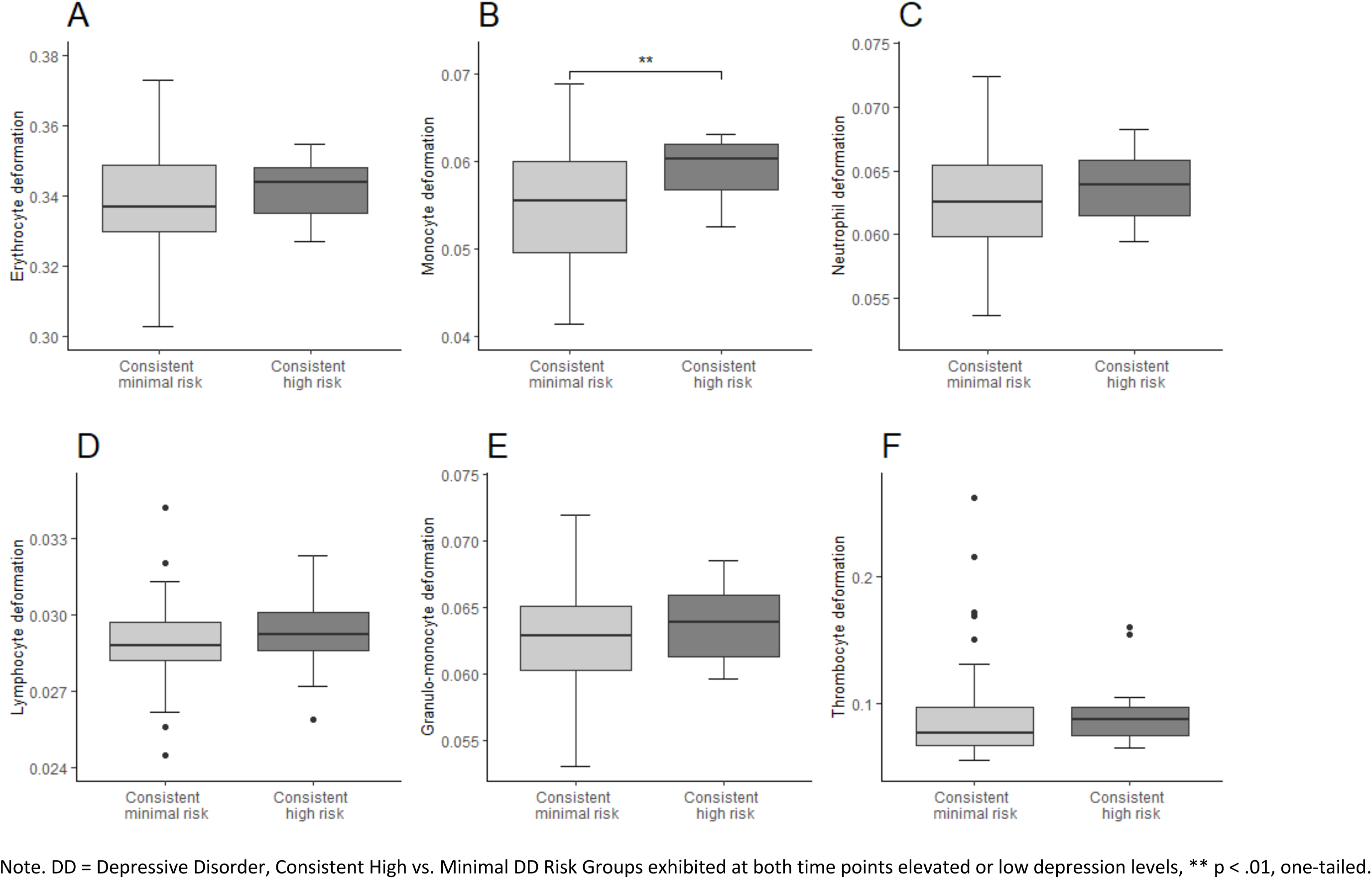
Combined Boxplots for the Consistent High DD Risk Group and the Consistent Minimal DD Risk Group Regarding Blood Cell Deformability

### Association of Cortisol and Cortisone *(T1 and T2)* with Immune Cell Count

Significant Pearson correlations emerged between cortisol and immune cell count in both PDD groups (see Supplementary Tables 14 and 15). Specifically, cortisol T1 was associated with the number of granulo-monocytes in the lifetime PDD (*r* = .29, *p* = .0377) and the 12-month PDD group (*r* = .52, *p* = .0035). Furthermore, in the 12-month PDD group, cortisol and cortisone at T1 were associated with neutrophil cell count (cortisol T1: *r* = .46, *p* = .0123, cortisone T1: *r* = .47, p = .0102), while cortisol T2 was associated with granulo-monocyte cell count (*r* = .3759, *p* = .04447) and cortisone T2 with neutrophil cell count (*r* = .38, .0416). Moreover, cortisone T1 and T2 were associated with the number of lymphocytes in the lifetime PDD group (T1: *r* = .3036, *p* = .0303; T2: *r* = .2758, *p* = .04996). However, only the association of cortisol and cortisone T1 with granulo-monocyte cell count remained significant in the 12-month PDD group after applying Holm-Bonferroni correction (cortisol T1: *p* = .0176; cortisone T1: *p* = .0153).

## Discussion

### Summary of Results

Depressive symptoms at T1 were positively associated with monocyte and lymphocyte deformability at T2 over the entire sample, while individuals with high depressive symptoms at T1 and consistently high depressive symptoms at both time points (T1 and T2) exhibited increased monocyte and lymphocyte deformability at T2 as compared to individuals with no or minimal depressive symptomatology. This pattern was most pronounced in the 12-month PDD and the lifetime PDD groups. An increase in depressive symptoms from T1 to T2 was not associated with cell deformability at T2. Basal glucocorticoid levels at T1 or T2 were further not associated with cell deformability at T2. Albeit not in the entire sample, in both the 12-month PDD and the lifetime PDD group, an increase in glucocorticoid secretion from T1 to T2 was associated with higher granulo-monocyte and neutrophil deformability. Similarly, higher glucocorticoid levels at T1 or T2 were associated with increased granulo-monocyte, neutrophil, and lymphocyte count in 12-month and lifetime PDD groups. Notably, there was no difference in cell size nor in immune cell counts between DD groups and healthy controls, nor was there an association between depressive symptoms and cell size. In addition, no difference in glucocorticoid levels was observed between DD groups and healthy controls, neither was medication intake associated with cell deformability.

### Integration of Findings

The present study is the first to examine the longitudinal relationship between depressive symptoms, glucocorticoid secretion and immune cell deformability in depressed individuals and healthy controls. The finding that elevated depressive symptomatology is associated with higher immune cell deformability one year later is supported by previously reported cross-sectional findings on the positive association between depressive disorder status, depressive symptoms and immune cell deformability (Walther et al., 2022). Because these supporting findings originate from the same study, it is important that further longitudinal as well as experimental studies replicate and extend these associations.

The observed significant correlations between depressive symptoms at T1 and immune cell deformability at T2 for monocytes and lymphocytes in the entire sample and for monocytes, neutrophils, and granulo-monocytes in the lifetime PDD and healthy control subsample suggests these immune cells to be the most sensitive cells to react to depressive symptomatology with physical changes. Since these cells are rich in glucocorticoid receptors (Goecke et al., 2007; Heiske et al., 2003; Li et al., 2006; Lu et al., 2017), and HPA-axis alterations are considered a pathophysiological landmark of DD (Rothe et al., 2020; Stetler and Miller, 2011), it is reasonable to assume that changes in glucocorticoid secretion might also affect immune cell status and deformability. Indeed, it was previously shown that increased glucocorticoid levels led to increased immune cell counts due to cell demargination from the vessel walls, which was mediated by cellular softening (Fay et al., 2016). However, in the present study, no direct association was identified between glucocorticoid levels at T1 or T2 with cell deformability at T2. Nonetheless, when examining the change in glucocorticoid secretion between T1 and T2, an increase in glucocorticoid secretion was associated with increased neutrophil and granulo-monocyte deformability at T2 in the lifetime and 12-month PDD and healthy control samples. Notably, we did not observe elevated levels of circulating immune cells in individuals suffering from DD in the present study (see Supplementary Table 6), although this was previously reported by another study (Lynall et al., 2019). In line with previous reports (Gerritsen et al., 2019; Rothe et al., 2020), no direct association between hair glucocorticoid levels and depressive disorder status emerged (see Supplementary Table 2 and 3). However, higher glucocorticoid levels were positively associated with immune cell count in the 12-month and lifetime PDD groups (see Supplementary Table 14 and 15), supporting the perspective that increased glucocorticoid levels support the demargination of immune cells from vessel walls mediated by cellular softening (Fay et al., 2016).

The results suggest that changes in glucocorticoid concentrations over a one-year period may affect immune cell structure and stability. Immune cells might respond to increased glucocorticoid concentrations by reorganizing the cytoskeleton, which affects cell deformability directly as previous studies showed using dexamethasone administration (Fay et al., 2016; Ronchetti et al., 2018). Moreover, elevated glucocorticoid concentrations can cause increased permeation through the cell membrane affecting the lipidome and the cell structure leading to membrane destabilization and bending (Walther et al., 2018), and hence to increased cell deformability. Alterations in membrane-forming lipids in the central nervous system have already been linked to DD (Müller et al., 2015).

The finding that 12-month and lifetime PDD show the strongest associations with cell deformability could be explained by the persistence of disease burden. Indeed, in analyses comparing those individuals with consistently high depressive symptoms (consistent high risk) with individuals with consistently low depressive symptoms (consistent minimal risk) regardless of the DD diagnosis at T2, we found further evidence for this perspective. Individuals exhibiting severely elevated depressive symptomatology at T1 as well as consistently severely elevated depressive symptomatology at both timepoints (T1 and T2) compared to individuals with no or minimal depressive symptoms, showed increased monocyte and lymphocyte deformability at T2. The results suggest that the time factor, namely suffering from severe depressive symptoms at least over a one-year period, might be central with regard to immune cell deformability in DD. Even though significant results can only be observed for monocytes and lymphocytes in the present analysis due to the rather small group sizes, it can be seen from Figure 2 and 3 that this tends to be the case for all immune cells. Given the persistence of depressive symptoms in PDD over two years, associations with immune cell deformability are more likely to be detected than in MDD, which only requires as persistence of symptomatology for over two weeks (APA, 2013). It is also likely that in PDD, the HPA axis shows hyperactivity with increased glucocorticoid output, as there is less depressive symptomatology but over a longer period of time than in MDD. In MDD, the HPA axis appears to enter exhaustion at some point, at which point a shift from increased to decreased levels of glucocorticoids are more likely (Rothe et al., 2020; Steudte-Schmiedgen et al., 2017, 2016; Walther et al., 2022a). Thus, many reports also identify reduced hair cortisol levels or more complex patterns in MDD and especially in conjunction with experienced trauma (Cantave et al., 2022; Hinkelmann et al., 2013; Psarraki et al., 2021; Stalder et al., 2017). Therefore, future studies examining the relationship between DD, glucocorticoid output and immune cell deformability should aim to assess the time period since when individuals are suffering from depressive symptomatology as well as childhood maltreatment or trauma experience to better disentangle these seemingly complex associations.

Overall, the present study provides new insights into the pathophysiology of DD, assuming more deformed immune cells, especially monocytes, with persistent depressive symptomatology. In addition to increased glucocorticoid output, research needs to investigate whether further processes consistently linked to DD such as low-grade inflammation (Goldsmith et al., 2016; Iob et al., 2020; Pariante, 2017) or oxidative stress (Moylan et al., 2014) might be additional factors contributing to cytoskeletal alterations and membrane destabilization (Wilson & González-Billault, 2015; Wong et al., 2013) resulting in higher immune cell deformability in DD. This might be because, similar to increased glucocorticoid secretion, an inflammatory state leads to demargination of blood cells from the vessel walls into the circulation, with cell deformability being increased by the migration process (Fay et al., 2016). Additionally, it was shown that an experimentally induced immune activation with lipopolysaccharide increased monocyte deformability (Ravetto et al., 2014), highlighting chronic low-grade inflammation in depressed individuals as potential agent for increased immune cell deformability. These assumptions are supported by findings showing that individuals with increased inflammatory signaling, such as healthy individuals after inhalation of lipopolysaccharide from E. coli, or individuals suffering from acute lung injury, viral respiratory infections, or Epstein-Barr virus infections, show increased deformability of neutrophils, monocytes, or lymphocytes (Toepfner et al., 2018a). Moreover, neutrophils of individuals in the acute phase of COVID-19 infection showed higher deformability, which was still observable also seven months after the acute infection symptoms (Kubánková et al., 2021), suggesting that long-term HPA-axis or inflammatory alterations may underlie immune cell deformability in DD.

It is important to mention that medication intake was not associated with immune cell deformability. As subjects were not drug naïve, we tested the association of different medication groups with cell deformability, such as psychopharmaceutical medication, antihypertensive drugs or thyroid dysfunction medication. No association between any medication group and cell deformability was identified. Also, associations between depressive symptomatology and cell deformability cannot be traced back to cell size, as higher depression severity and cell size of immune cells were not associated with each other, nor did cell size differ between groups with DD and matched healthy controls.

### Strengths and Limitations

Some limitations must be taken into account when interpreting the results. The most important limitation is the single measurement of cell deformability at T2. To investigate the predictive value of depressive symptoms, glucocorticoid concentrations as well as their changes over a one-year period on cell deformability, we measured cell deformability as an outcome measure at T2. However, analyses of changes in cell deformability in relation to changes in depressive symptoms or glucocorticoid concentrations would also be important to investigate. Furthermore, with such a study design, the overall stability of cell deformability markers could be recorded. A related point is the single assessment of the diagnostic groups by clinical interview at T2. Since we intended to capture group differences in cell deformability (Walther et al., 2022b), the clinical diagnostics as well as the cell deformability measurement had to be performed at the same time point in order to avoid possible time effects. However, in a follow-up project, clinical diagnostics will also be performed at two time points together with cell deformability measurement. Nevertheless, the present findings show for the first time that depressive symptoms are related to immune cell deformability one year later, whereas the increase in glucocorticoid secretion over one year is also associated with increased immune cell deformability. Following on from this, however, it should be noted that other important systemic markers have not been investigated in the present study. Thus, it will be important in future studies to explore potential associations between inflammatory markers such as Interleukin-6 or C-reactive protein and immune cell deformability. In addition, the present sample of 136 participants might be underpowered to detect small to moderate effects, so only relatively large effects were probably identified in the present study further supporting the strong relationship between depressive symptoms, glucocorticoid secretion and immune cell deformability. Finally, gender differences could also play a role not revealed in this study, so that larger mixed samples or samples that focus only on women, men, or gender-diverse individuals need to be further investigated.

### Conclusions

Taken together, this study represents the first longitudinal investigation of the association between depressive symptoms, glucocorticoid concentrations, and peripheral immune cell deformability in individuals with DD and healthy controls. Our results suggest that higher levels of depressive symptoms are longitudinally associated with higher immune cell deformability, particularly in monocytes and lymphocytes. Moreover, we observed partial confirmation for the hypothesis that DD-related increases in glucocorticoid concentrations lead to cytoskeletal changes and cell membrane destabilization causing overall increased immune cell deformability. Increased immune cell deformability may further lead to a gradual loss of immune cell functionality, so that, for example, immune response is impaired or membrane function is compromised presenting a potentially underlying mechanism causing or maintaining DD.

## Contributors

AW, JE and MK designed the study, performed experiments and data analysis. AW wrote the first draft of the manuscript and and JE wrote the statistical analysis and results section of the manuscript. MK, WG, MW, MP, NR performed experiments. CK, JG, MK, LW provided funding, infrastructure/equipment, and edited subsequent versions of the manuscript.

## Declaration of interest

All authors state that they have no actual or potential conflict of interest to declare, including any financial, personal, or other relationships with other people or organisations within 3 years of beginning the submitted work that could influence or bias their work.

### Data and code availability

The anonymized data and code will be made available to all interested parties upon request.

## Data Availability

All data produced in the present study are available upon reasonable request to the authors

## Supplementary Material

**Supplementary Table 2.**
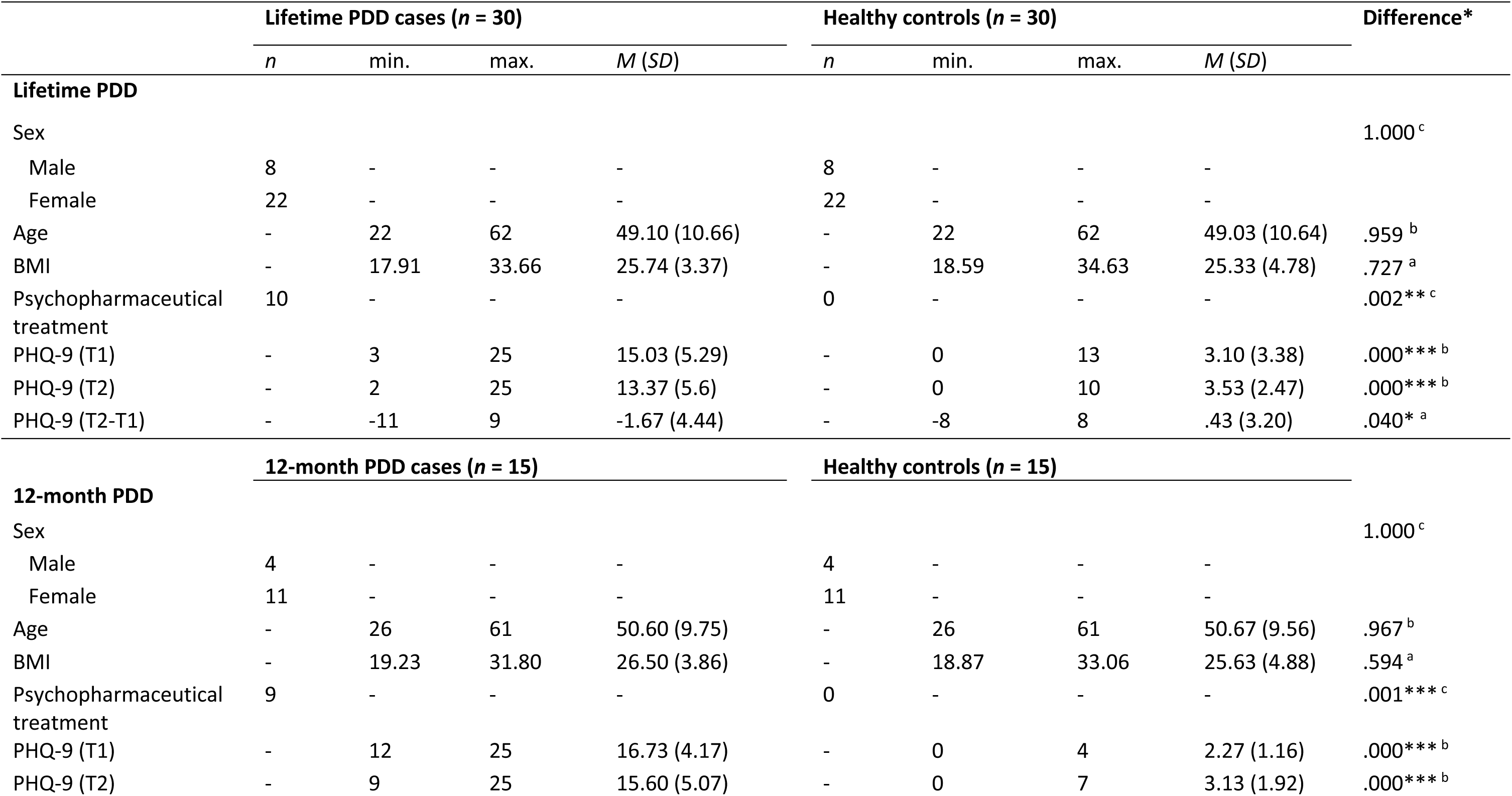
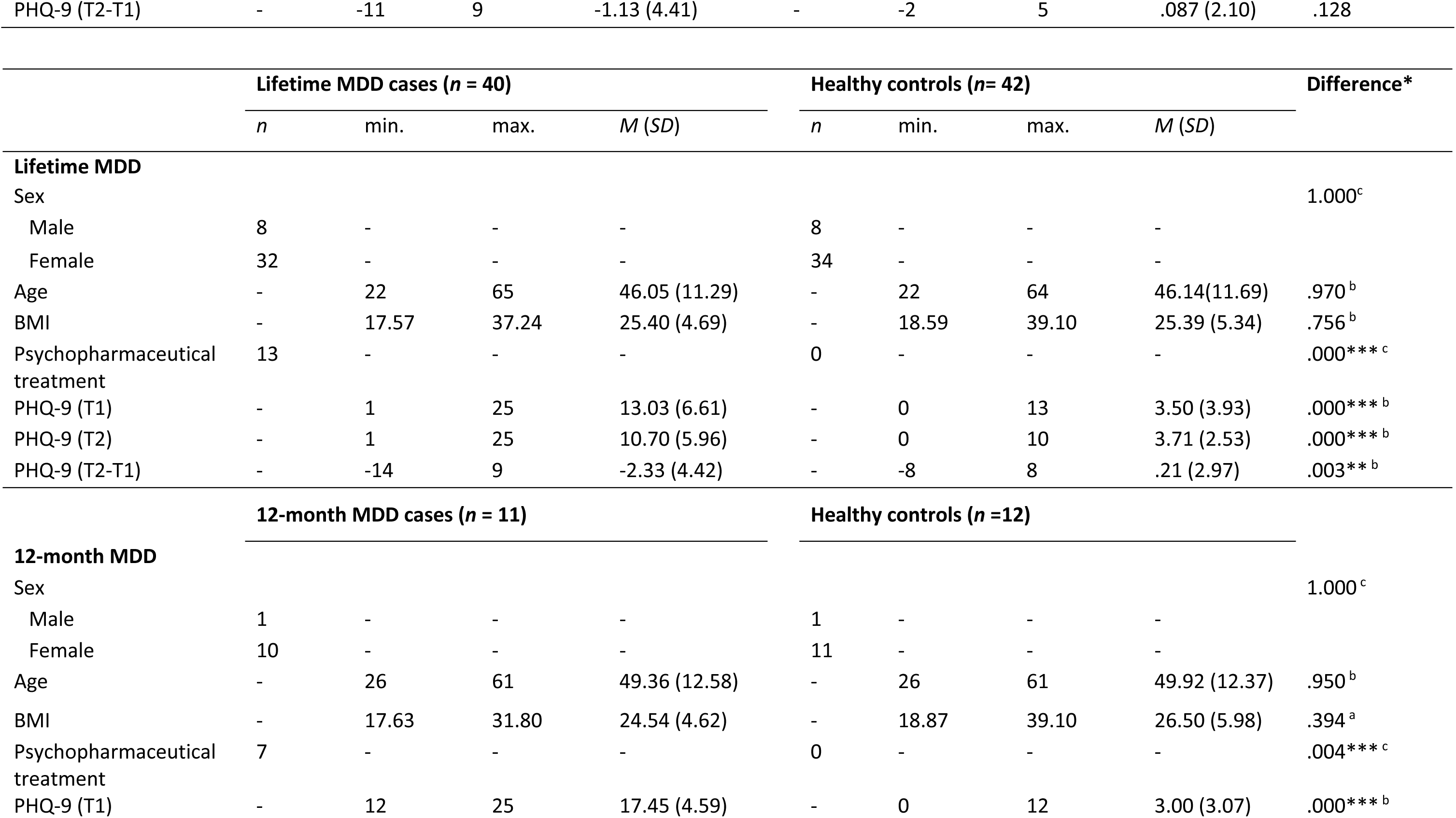
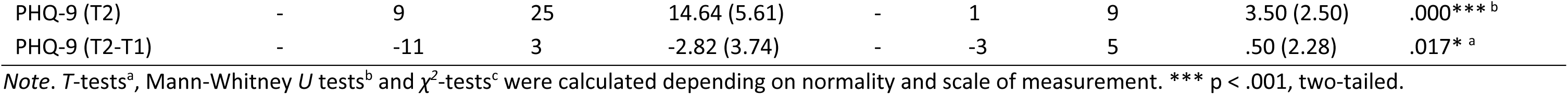
Sample Characteristics According to Different Diagnostic Groups

**Supplementary Table 3.**
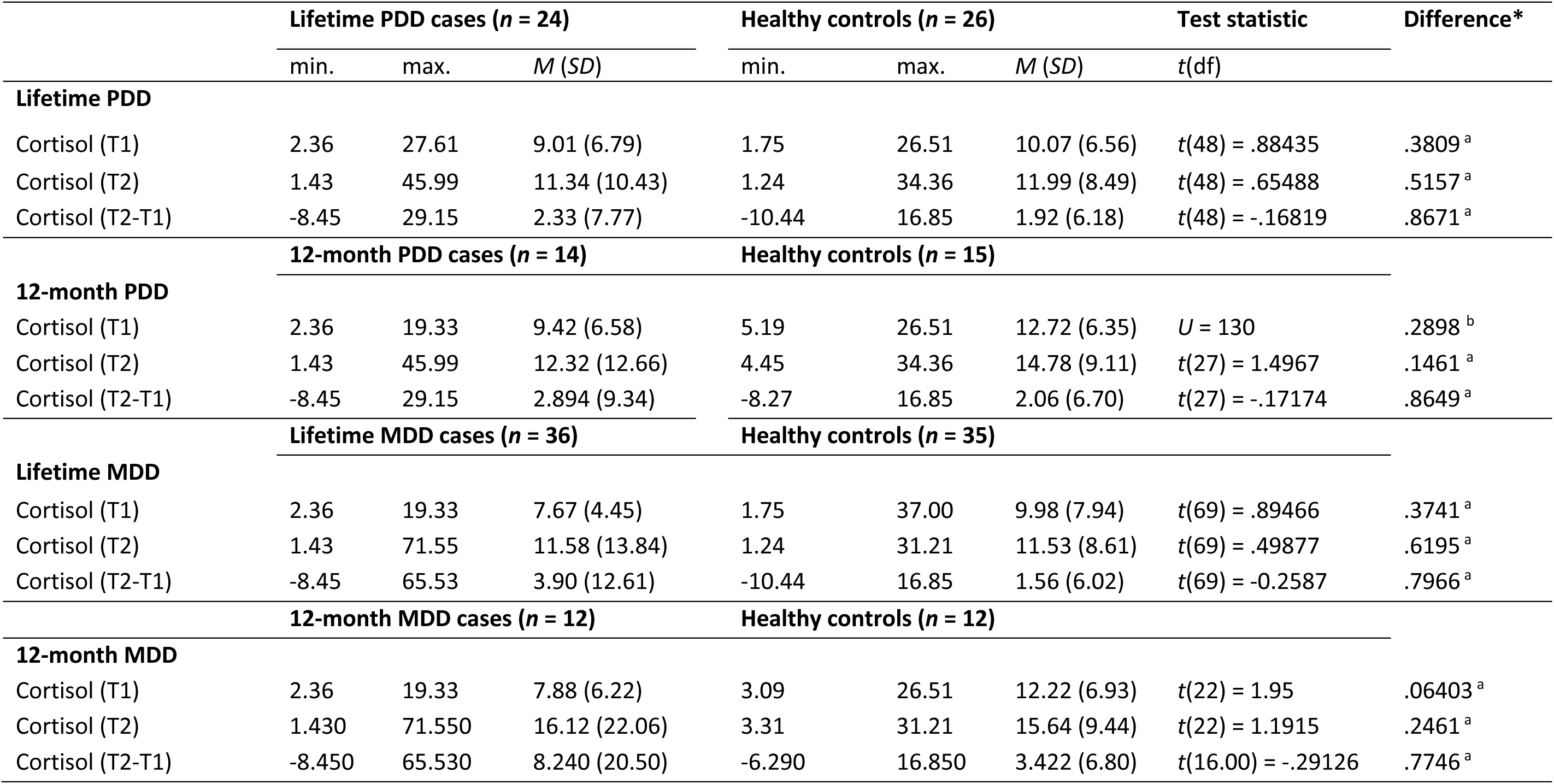
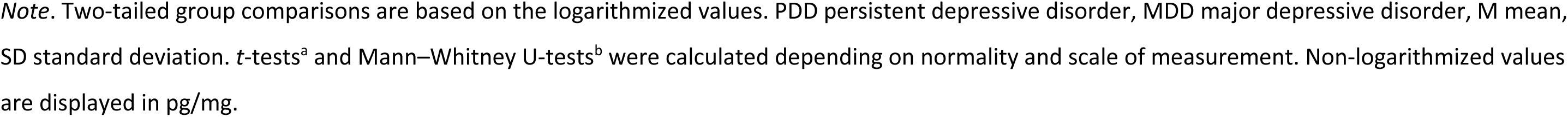
Mean Group Comparisons of Cortisol Between Different Diagnostic Groups and Healthy Controls

**Supplementary Table 4.**
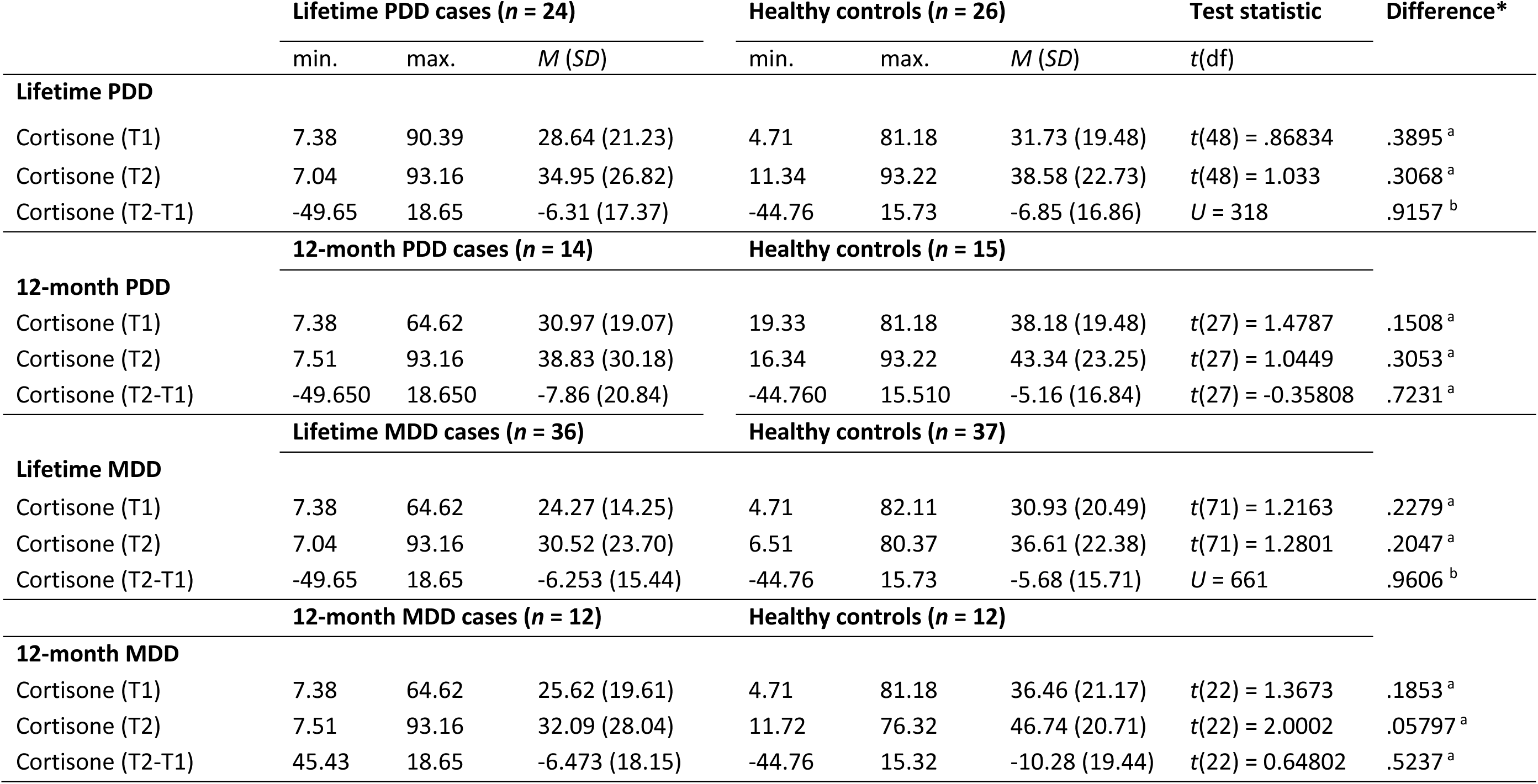
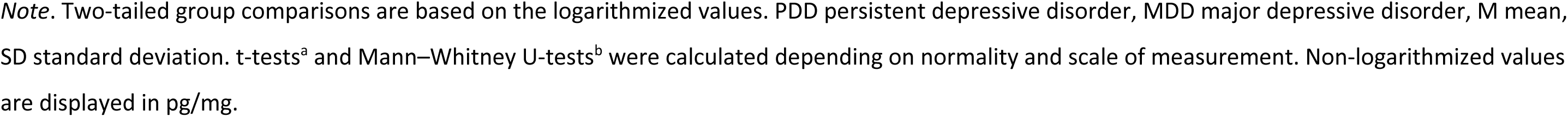
Mean Group Comparisons of Cortisone Between Different Diagnostic Groups and Healthy Controls

|  | **Lifetime PDD cases (*n* = 24)** |  |  | **Healthy controls (*n* = 26)** |  |  | **Test statistic** | **Difference\*** |
| --- | --- | --- | --- | --- | --- | --- | --- | --- |
|  | min. | max. | *M* (*SD*) | min. | max. | *M* (*SD*) | *t*(*df*) |  |
| **Lifetime PDD** |  |  |  |  |  |  |  |  |
| Cortisone (T1) | 7.38 | 90.39 | 28.64 (21.23) | 4.71 | 81.18 | 31.73 (19.48) | *t*(48) = .86834 | .3895<sup>a</sup> |
| Cortisone (T2) | 7.04 | 93.16 | 34.95 (26.82) | 11.34 | 93.22 | 38.58 (22.73) | *t*(48) = 1.033 | .3068<sup>a</sup> |
| Cortisone (T2-T1) | -49.65 | 18.65 | -6.31 (17.37) | -44.76 | 15.73 | -6.85 (16.86) | *U* = 318 | .9157<sup>b</sup> |
| **12-month PDD** |  |  |  |  |  |  |  |  |
|  | **12-month PDD cases (*n* = 14)** |  |  | **Healthy controls (*n* = 15)** |  |  |  |  |
| Cortisone (T1) | 7.38 | 64.62 | 30.97 (19.07) | 19.33 | 81.18 | 38.18 (19.48) | *t*(27) = 1.4787 | .1508<sup>a</sup> |
| Cortisone (T2) | 7.51 | 93.16 | 38.83 (30.18) | 16.34 | 93.22 | 43.34 (23.25) | *t*(27) = 1.0449 | .3053<sup>a</sup> |
| Cortisone (T2-T1) | -49.650 | 18.650 | -7.86 (20.84) | -44.760 | 15.510 | -5.16 (16.84) | *t*(27) = -0.35808 | .7231<sup>a</sup> |
| **Lifetime MDD** |  |  |  |  |  |  |  |  |
|  | **Lifetime MDD cases (*n* = 36)** |  |  | **Healthy controls (*n* = 37)** |  |  |  |  |
| Cortisone (T1) | 7.38 | 64.62 | 24.27 (14.25) | 4.71 | 82.11 | 30.93 (20.49) | *t*(71) = 1.2163 | .2279<sup>a</sup> |
| Cortisone (T2) | 7.04 | 93.16 | 30.52 (23.70) | 6.51 | 80.37 | 36.61 (22.38) | *t*(71) = 1.2801 | .2047<sup>a</sup> |
| Cortisone (T2-T1) | -49.65 | 18.65 | -6.253 (15.44) | -44.76 | 15.73 | -5.68 (15.71) | *U* = 661 | .9606<sup>b</sup> |
| **12-month MDD** |  |  |  |  |  |  |  |  |
|  | **12-month MDD cases (*n* = 12)** |  |  | **Healthy controls (*n* = 12)** |  |  |  |  |
| Cortisone (T1) | 7.38 | 64.62 | 25.62 (19.61) | 4.71 | 81.18 | 36.46 (21.17) | *t*(22) = 1.3673 | .1853<sup>a</sup> |
| Cortisone (T2) | 7.51 | 93.16 | 32.09 (28.04) | 11.72 | 76.32 | 46.74 (20.71) | *t*(22) = 2.0002 | .05797<sup>a</sup> |
| Cortisone (T2-T1) | 45.43 | 18.65 | -6.473 (18.15) | -44.76 | 15.32 | -10.28 (19.44) | *t*(22) = 0.64802 | .5237<sup>a</sup> |

*Note.* Two-tailed group comparisons are based on the logarithmized values. PDD persistent depressive disorder, MDD major depressive disorder, M mean, SD standard deviation. t-tests<sup>a</sup> and Mann–Whitney U-tests<sup>b</sup> were calculated depending on normality and scale of measurement. Non-logarithmized values are displayed in pg/mg.

**Supplementary Table 4.** Associations Between PHQ-9 (T1) and Cell Area According to the Entire Sample

| **Cell Area** | **Erythrocytes** | **Monocytes** | **Neutrophils** | **Lymphocytes** | **Granulo-Monocytes** | **Thrombocytes** |
| --- | --- | --- | --- | --- | --- | --- |
| Pearson | -.0302<br>*p* = .6363 | .1297<br>*p* = .066 | .0667<br>*p* = .2202 | -.0760<br>*p* = .7929 | .0261<br>*p* = .3816 | -.0478<br>*p* = .7099 |
| Kendall | -.0508<br>*p* = .7995 | .0804<br>*p* = .0913 | .0598<br>*p* = .1617 | -.0275<br>*p* = .6744 | .0415<br>*p* = .2468 | -.0003<br>*p* = .5023 |
| Partial Correlation | -.0468<br>*p* = .7030 | .1350<br>*p* = .0614 | .0559<br>*p* = .2622 | -.0930<br>*p* = .8562 | .0181<br>*p* = .4169 | -.0188<br>*p* = .5849 |

*Note.* Pearson and Kendall tests were calculated depending on normality and scale of measurement (*n* = 136). All correlations were calculated one-tailed.

**Supplement Table 5.** Associations Between PHQ-9 (T2) and Cell Area According to the Entire Sample

| **Cell Area** | **Erythrocytes** | **Monocytes** | **Neutrophils** | **Lymphocytes** | **Granulo-Monocytes** | **Thrombocytes** |
| --- | --- | --- | --- | --- | --- | --- |
| Pearson | -.0145<br>*p* = .5664 | .0637<br>*p* = .2305 | .0207<br>*p* = .4055 | -.0519<br>*p* = .7258 | -.0194<br>*p* = .5886 | -.0341<br>*p* = .6532 |
| Kendall | -.0378<br>*p* = .7358 | .0315<br>*p* = .2987 | .0250<br>*p* = .3383 | -.0328<br>*p* = .7073 | .0130<br>*p* = .4141 | -.0367<br>*p* = .7317 |
| Partial<br>Correlation | -.0247<br>*p* = .6108 | .0708<br>*p* = .2100 | .0063<br>*p* = .4716 | -.0713<br>*p* = .7917 | -.0297<br>*p* = .6324 | -.0100<br>*p* = .5453 |

*Note.* Pearson and Kendall tests were calculated depending on normality and scale of measurement (*n* = 136). All correlations were calculated one-tailed.

**Supplementary Table 6.** Mean Difference of Immune Cell Count Between Different Diagnostic Groups and Matched Healthy Controls

|  | **Monocytes** | **Neutrophils** | **Lymphocytes** | **Granulo-Monocytes** |
| --- | --- | --- | --- | --- |
| **Lifetime PDD (*n* = 30)** |  |  |  |  |
| M | 41.87 | 340.63 | 265.37 | 402.97 |
| SD | 65.52 | 174.68 | 98.82 | 184.87 |
| **HC (*n* = 30)** |  |  |  |  |
| M | 33.83 | 365.43 | 292.47 | 419.03 |
| SD | 34.02 | 218.10 | 116.45 | 222.89 |
| *U*-value | *U* = 454 | *U* = 484 | *U* = 517.5 | *U* = 486 |
| *p*-value | .9561 | .6202 | .3326 | .7945 |
| **12-month PDD (*n* = 15)** |  |  |  |  |
| M | 59.67 | 358.13 | 258.6 | 443.33 |
| SD | 90.04 | 211.45 | 106.06 | 221.31 |
| **HC (*n* = 15)** |  |  |  |  |
| M | 39.80 | 344.07 | 300.0 | 404.33 |
| SD | 44.96 | 162.83 | 110.19 | 171.00 |
| *U*-value | *U* = 107 | *U* = 116 | *U* = 138 | *U* = 103.5 |
| *p*-value | .8301 | .9025 | .3046 | .7209 |
| **Lifetime MDD (*n* = 40)** |  |  |  |  |
| M | 33.95 | 350.00 | 272.40 | 406.73 |
| SD | 47.63 | 172.61 | 135.04 | 185.56 |
| **HC (*n* = 42)** |  |  |  |  |
| M | 44.36 | 360.76 | 316.00 | 428.81 |
| SD | 74.40 | 203.00 | 223.25 | 208.53 |
| *U*-value | *U* = 911.5 | *U* = 835 | *U* = 934 | *U* = 869 |
| *p*-value | .5105 | .965 | .3864 | .7908 |

|  | Monocytes | Neutrophils | Lymphocytes | Granulo-Monocytes |
| --- | --- | --- | --- | --- |
| **12-month MDD (*n* = 11)** |  |  |  |  |
| M | 30.36 | 369.36 | 254.64 | 427.45 |
| SD | 16.98 | 152.41 | 118.42 | 185.48 |
| **HC (*n* = 12)** |  |  |  |  |
| M | 44.83 | 349.83 | 320.75 | 424.58 |
| SD | 45.48 | 195.10 | 122.52 | 208.17 |
| *U*-value | *U* = 77 | *U* = 59 | *U* = 85 | *U* = 69.5 |
| *p*-value | .5155 | .6947 | .2604 | .8450 |

*Note.* Two-tailed *t*-tests and Mann-Whitney *U* tests were calculated depending on normality and scale of measurement.

**Supplementary Table 7.** Associations Between Depressive Symptoms Measured by PHQ-9 (T2) and Cell Deformation Over the Entire Sample

|  | **Erythrocytes** | **Monocytes** | **Neutrophils** | **Lymphocytes** | **Granulo-**<br>**monocytes** | **Thrombocytes** |
| --- | --- | --- | --- | --- | --- | --- |
| Pearson | -.0529<br>*p* = .7296 | .1678*<br>*p* = .0254 | .0588<br>*p* = .2481 | .0843<br>*p* = .1645 | .0584<br>*p* = .2497 | .0644<br>*p* = .2281 |
| Kendall | -.0224<br>*p* = .6457 | .1139*<br>*p* = .0278 | .0426<br>*p* = .2373 | .0542<br>*p* = .1825 | .0437<br>*p* = .2315 | .0690<br>*p* = .1230 |
| Partial<br>correlation | -.0403<br>*p* = .6770 | .1567*<br>*p* = .0363 | .0074<br>*p* = .4663 | .0881<br>*p* = .1577 | .0069<br>*p* = .4689 | .0498<br>*p* = .2854 |

*Note.* Pearson and Kendall correlations were calculated depending on linearity and scale of measurement. All correlations were calculated one-tailed. \* *p* < .05, one-tailed. \*\* *p* < .01, one-tailed.

**Supplementary Table 8.**
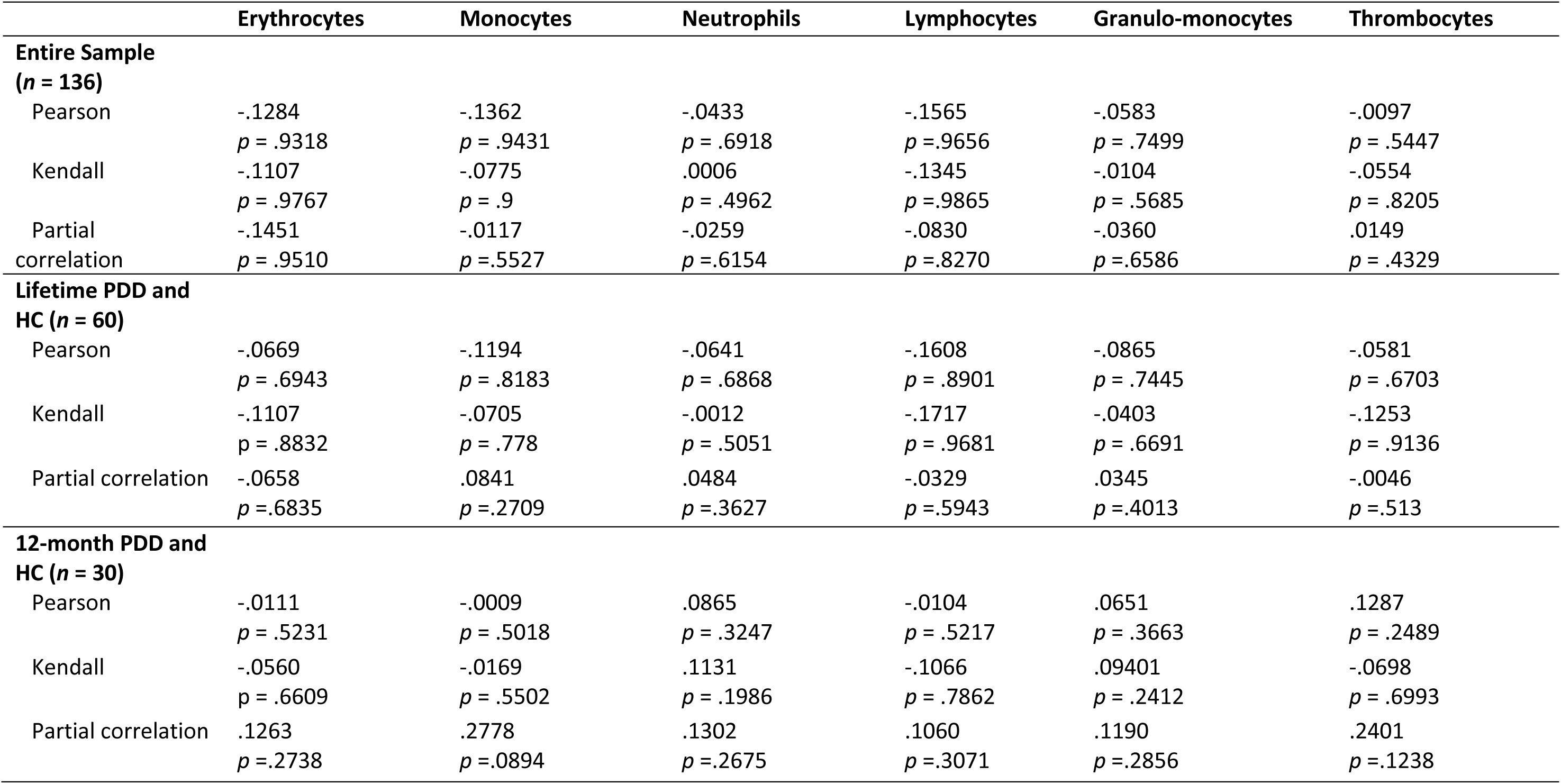
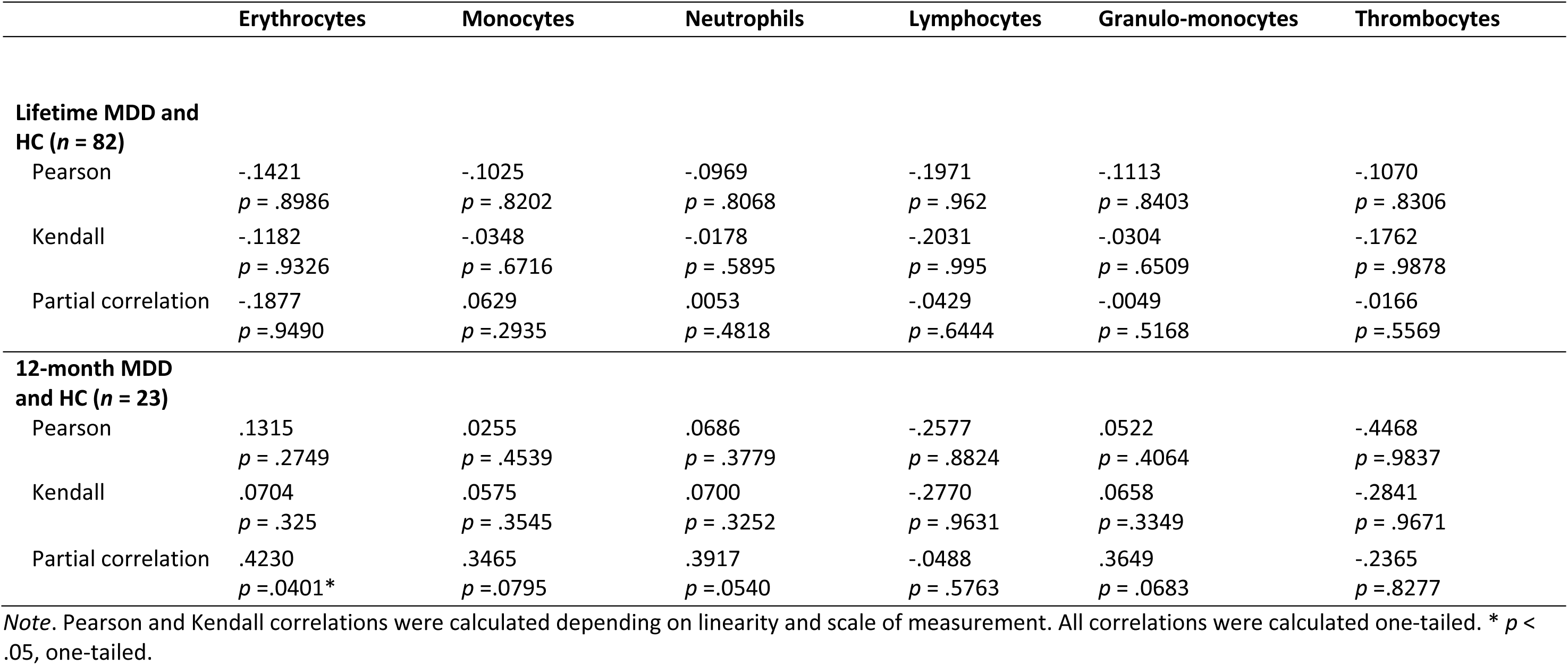
Associations Between Depressive Symptoms Difference Measured by PHQ-9 (T2-T1) and Cell Deformability Over the Entire Sample and According to Different Diagnostic Groups

|  | Erythrocytes | Monocytes | Neutrophils | Lymphocytes | Granulo-monocytes | Thrombocytes |
| --- | --- | --- | --- | --- | --- | --- |
| **Entire Sample**<br>**(n = 136)** |  |  |  |  |  |  |
| Pearson | -.1284<br>*p* = .9318 | -.1362<br>*p* = .9431 | -.0433<br>*p* = .6918 | -.1565<br>*p* = .9656 | -.0583<br>*p* = .7499 | -.0097<br>*p* = .5447 |
| Kendall | -.1107<br>*p* = .9767 | -.0775<br>*p* = .9 | .0006<br>*p* = .4962 | -.1345<br>*p* = .9865 | -.0104<br>*p* = .5685 | -.0554<br>*p* = .8205 |
| Partial correlation | -.1451<br>*p* = .9510 | -.0117<br>*p* = .5527 | -.0259<br>*p* = .6154 | -.0830<br>*p* = .8270 | -.0360<br>*p* = .6586 | .0149<br>*p* = .4329 |
| **Lifetime PDD and**<br>**HC (n = 60)** |  |  |  |  |  |  |
| Pearson | -.0669<br>*p* = .6943 | -.1194<br>*p* = .8183 | -.0641<br>*p* = .6868 | -.1608<br>*p* = .8901 | -.0865<br>*p* = .7445 | -.0581<br>*p* = .6703 |
| Kendall | -.1107<br>*p* = .8832 | -.0705<br>*p* = .778 | -.0012<br>*p* = .5051 | -.1717<br>*p* = .9681 | -.0403<br>*p* = .6691 | -.1253<br>*p* = .9136 |
| Partial correlation | -.0658<br>*p* = .6835 | .0841<br>*p* = .2709 | .0484<br>*p* = .3627 | -.0329<br>*p* = .5943 | .0345<br>*p* = .4013 | -.0046<br>*p* = .513 |
| **12-month PDD and**<br>**HC (n = 30)** |  |  |  |  |  |  |
| Pearson | -.0111<br>*p* = .5231 | -.0009<br>*p* = .5018 | .0865<br>*p* = .3247 | -.0104<br>*p* = .5217 | .0651<br>*p* = .3663 | .1287<br>*p* = .2489 |
| Kendall | -.0560<br>*p* = .6609 | -.0169<br>*p* = .5502 | .1131<br>*p* = .1986 | -.1066<br>*p* = .7862 | .09401<br>*p* = .2412 | -.0698<br>*p* = .6993 |
| Partial correlation | .1263<br>*p* = .2738 | .2778<br>*p* = .0894 | .1302<br>*p* = .2675 | .1060<br>*p* = .3071 | .1190<br>*p* = .2856 | .2401<br>*p* = .1238 |
| **Lifetime MDD and HC (*n* = 82)** |  |  |  |  |  |  |
| Pearson | -.1421<br>*p* = .8986 | -.1025<br>*p* = .8202 | -.0969<br>*p* = .8068 | -.1971<br>*p* = .962 | -.1113<br>*p* = .8403 | -.1070<br>*p* = .8306 |
| Kendall | -.1182<br>*p* = .9326 | -.0348<br>*p* = .6716 | -.0178<br>*p* = .5895 | -.2031<br>*p* = .995 | -.0304<br>*p* = .6509 | -.1762<br>*p* = .9878 |
| Partial correlation | -.1877<br>*p* = .9490 | .0629<br>*p* = .2935 | .0053<br>*p* = .4818 | -.0429<br>*p* = .6444 | -.0049<br>*p* = .5168 | -.0166<br>*p* = .5569 |
| **12-month MDD and HC (*n* = 23)** |  |  |  |  |  |  |
| Pearson | .1315<br>*p* = .2749 | .0255<br>*p* = .4539 | .0686<br>*p* = .3779 | -.2577<br>*p* = .8824 | .0522<br>*p* = .4064 | -.4468<br>*p* = .9837 |
| Kendall | .0704<br>*p* = .325 | .0575<br>*p* = .3545 | .0700<br>*p* = .3252 | -.2770<br>*p* = .9631 | .0658<br>*p* = .3349 | -.2841<br>*p* = .9671 |
| Partial correlation | .4230<br>*p* = .0401* | .3465<br>*p* = .0795 | .3917<br>*p* = .0540 | -.0488<br>*p* = .5763 | .3649<br>*p* = .0683 | -.2365<br>*p* = .8277 |

*Note.* Pearson and Kendall correlations were calculated depending on linearity and scale of measurement. All correlations were calculated one-tailed. \* *p* < .05, one-tailed.

**Supplementary Table 9.** Associations Between Cortisol T1 and Cell Deformability Over the Entire Sample and According to Different Diagnostic Groups

|  | Erythrocytes | Monocytes | Neutrophils | Lymphocytes | Granulo-monocytes | Thrombocytes |
| --- | --- | --- | --- | --- | --- | --- |
| **Entire Sample**<br>**(n = 116)** |  |  |  |  |  |  |
| Pearson | -.1198<br>*p* = .2002 | -.0784<br>*p* = .4027 | .1155<br>*p* = .2168 | .1722<sup>†</sup><br>*p* = .0645 | .0858<br>*p* = .3600 | .1243<br>*p* = .1836 |
| Kendall | -.0825<br>*p* = .1932 | -.0534<br>*p* = .3969 | .0643<br>*p* = .3082 | .0801<br>*p* = .2066 | .0541<br>*p* = .3916 | .0455<br>*p* = .4697 |
| Partial correlation | -.0665<br>*p* = .4861 | -.0821<br>*p* = .3897 | .0958<br>*p* = .3150 | .0927<br>*p* = .3309 | .0610<br>*p* = .5232 | .0995<br>*p* = .2967 |
| **Lifetime PDD and**<br>**HC (n = 50)** |  |  |  |  |  |  |
| Pearson | -.1232<br>*p* = .3938 | -.0382<br>*p* = .7922 | -.2232<br>*p* = .1193 | .2103<br>*p* = .1427 | -.2099<br>*p* = .1434 | .0577<br>*p* = .6905 |
| Kendall | -.1265<br>*p* = .200 | .0025<br>*p* = .9800 | -.1477<br>*p* = .1320 | .0692<br>*p* = .4819 | -.0959<br>*p* = .3276 | -.0498<br>*p* = .6098 |
| Partial correlation | -.1511<br>*p* = .3160 | .0164<br>*p* = .9139 | -.2509<sup>†</sup><br>*p* = .0925 | .1912<br>*p* = .2031 | -.2330<br>*p* = .1191 | .0170<br>*p* = .9105 |
| **12-month PDD and**<br>**HC (n = 29)** |  |  |  |  |  |  |
| Pearson | -.1942<br>*p* = .3128 | -.1266<br>*p* = .5128 | -.3219<br>*p* = .0886 | .1413<br>*p* = .4648 | -.3576<sup>†</sup><br>*p* = .0568 | -.0465<br>*p* = .8107 |
| Kendall | -.1446<br>*p* = .2756 | -.0321<br>*p* = .8072 | -.2343<br>*p* = .0747 | .0174<br>*p* = .8954 | -.2497<sup>†</sup><br>*p* = .0580 | -.1652<br>*p* = .2087 |
| Partial correlation | -.1956<br>*p* = .3488 | .0197<br>*p* = .9255 | -.3160<br>*p* = .1238 | .1994<br>*p* = .3393 | -.3418<sup>†</sup><br>*p* = .0945 | -.0230<br>*p* = .9129 |
| **Lifetime MDD and HC (n = 71)** |  |  |  |  |  |  |
| Pearson | -.0615<br>*p* = .6103 | -.1016<br>*p* = .3990 | .0195<br>*p* = .8719 | .1168<br>*p* = .3321 | -.0123<br>*p* = .9191 | .1557<br>*p* = .1949 |
| Kendall | -.0480<br>*p* = .5577 | -.0650<br>*p* = .4241 | -.0303<br>*p* = .7096 | .0556<br>*p* = .4962 | -.0267<br>*p* = .7431 | .0734<br>*p* = .3663 |
| Partial correlation | -.0249<br>*p* = .8418 | -.0882<br>*p* = .4778 | .0458<br>*p* = .7131 | .0942<br>*p* = .4483 | .0086<br>*p* = .9451 | .1146<br>*p* = .3558 |
| **12-month MDD and HC (n = 24)** |  |  |  |  |  |  |
| Pearson | -.1395<br>*p* = .5156 | -.2610<br>*p* = .2180 | -.1097<br>*p* = .6100 | -.0538<br>*p* = .8029 | -.1420<br>*p* = .5081 | -.0543<br>*p* = .8011 |
| Kendall | -.1241<br>*p* = .3984 | -.1561<br>*p* = .2860 | -.1416<br>*p* = .3332 | -.0949<br>*p* = .5185 | -.1232<br>*p* = .4172 | -.0399<br>*p* = .7849 |
| Partial correlation | -.1556<br>*p* = .5125 | -.2311<br>*p* = .3269 | -.1548<br>*p* = .5146 | -.1212<br>*p* = .6107 | -.1745<br>*p* = .4618 | -.0949<br>*p* = .6906 |

*Note.* Pearson and Kendall tests were calculated depending on normality and scale of measurement. All correlations were calculated two-tailed. <sup>†</sup> *p* < .1, two-tailed (trend).

**Supplementary Table 10.** Associations Between Cortisol T2 and Cell Deformability Over the Entire Sample and According to Different Diagnostic Groups

|  | Erythrocytes | Monocytes | Neutrophils | Lymphocytes | Granulo-monocytes | Thrombocytes |
| --- | --- | --- | --- | --- | --- | --- |
| **Entire Sample**<br>**(n = 116)** |  |  |  |  |  |  |
| Pearson | -.0470<br>*p* = .6162 | -.0172<br>*p* = .8545 | .0727<br>*p* = .4379 | .0294<br>*p* = .7544 | .0473<br>*p* = .6143 | -.0317<br>*p* = .7353 |
| Kendall | -.0835<br>*p* = .1876 | -.0036<br>*p* = .9543 | .0367<br>*p* = .5604 | .0109<br>*p* = .8635 | .0349<br>*p* = .5798 | -.0663<br>*p* = .2916 |
| Partial correlation | .0055<br>*p* = .9538 | -.0184<br>*p* = .8475 | .0524<br>*p* = .5830 | -.0507<br>*p* = .5952 | .0230<br>*p* = .8095 | -.0492<br>*p* = .6067 |
| **Lifetime PDD and**<br>**HC (n = 50)** |  |  |  |  |  |  |
| Pearson | -.1827<br>*p* = .2040 | .0202<br>*p* = .8894 | .0105<br>*p* = .9423 | .2255<br>*p* = .1154 | .0205<br>*p* = .8876 | .0641<br>*p* = .6582 |
| Kendall | -.1413<br>*p* = .1520 | .0451<br>*p* = .6453 | .0394<br>*p* = .6879 | .0922<br>*p* = .3484 | .0861<br>*p* = .3796 | -.009<br>*p* = .9267 |
| Partial correlation | -.2122<br>*p* = .1569 | .04431<br>*p* = .7700 | .0142<br>*p* = .9253 | .1991<br>*p* = .1846 | .0218<br>*p* = .8877 | .0619<br>*p* = .6826 |
| **12-month PDD and**<br>**HC (n = 29)** |  |  |  |  |  |  |
| Pearson | -.3763*<br>*p* = .0442 | -.0349<br>*p* = .8573 | -.0241<br>*p* = .9013 | .2946<br>*p* = .1208 | -.0322<br>*p* = .8682 | -.0152<br>*p* = .9377 |
| Kendall | -.2394<sup>†</sup><br>*p* = .0712 | -.0124<br>*p* = .9252 | .0271<br>*p* = .8365 | .0870<br>*p* = .5109 | .0420<br>*p* = .7497 | -.0617<br>*p* = .6390 |
| Partial correlation | -.3912<sup>†</sup><br>*p* = .0531 | -.0186<br>*p* = .9297 | .0390<br>*p* = .8533 | .2981<br>*p* = .1478 | .0294<br>*p* = .8892 | .0109<br>*p* = .9589 |
| **Lifetime MDD and HC (n = 71)** |  |  |  |  |  |  |
| Pearson | .0283<br>*p* = .8146 | -.0772<br>*p* = .5221 | -.0116<br>*p* = .9237 | -.0228<br>*p* = .8506 | -.0432<br>*p* = .7207 | .1007<br>*p* = .4033 |
| Kendall | -.0256<br>*p* = .7543 | -.0500<br>*p* = .5382 | -.0259<br>*p* = .7507 | -.0089<br>*p* = .9130 | -.0182<br>*p* = .8232 | .0415<br>*p* = .6091 |
| Partial correlation | .0637<br>*p* = .6085 | -.0491<br>*p* = .6930 | .0263<br>*p* = .8325 | -.0451<br>*p* = .7173 | -.0087<br>*p* = .9446 | .1239<br>*p* = .3178 |
| **12-month MDD and HC (n = 24)** |  |  |  |  |  |  |
| Pearson | -.0807<br>*p* = .7078 | .0896<br>*p* = .6770 | .0915<br>*p* = .6706 | .0395<br>*p* = .8546 | .0950<br>*p* = .6588 | .0641<br>*p* = .7661 |
| Kendall | -.1095<br>*p* = .4561 | .0327<br>*p* = .8233 | .0181<br>*p* = .9013 | -.0876<br>*p* = .5511 | .0507<br>*p* = .7498 | .0181<br>*p* = .9013 |
| Partial correlation | .0175<br>*p* = .9416 | .0276<br>*p* = .9079 | .0774<br>*p* = .7456 | -.0804<br>*p* = .7363 | .0705<br>*p* = .7678 | -.0305<br>*p* = .8984 |

*Note.* Pearson and Kendall tests were calculated depending on normality and scale of measurement. All correlations were calculated two-tailed. \* $p < .05$ , two-tailed. <sup>†</sup> $p < .1$ , two-tailed (trend).

**Supplementary Table 11.** Associations Between Cortisone T1 and Cell Deformability Over the Entire Sample and According to Different Diagnostic Groups

|  | Erythrocytes | Monocytes | Neutrophils | Lymphocytes | Granulo-monocytes | Thrombocytes |
| --- | --- | --- | --- | --- | --- | --- |
| **Entire Sample**<br>**(n = 118)** |  |  |  |  |  |  |
| Pearson | -.0673<br>*p* = .4692 | -.0131<br>*p* = .8884 | .1415<br>*p* = .1265 | .1550<sup>†</sup><br>*p* = .0937 | .1330<br>*p* = .1511 | .1175<br>*p* = .2051 |
| Kendall | -.0702<br>*p* = .2640 | -.0065<br>*p* = .9166 | .0619<br>*p* = .3217 | .0534<br>*p* = .3957 | .0646<br>*p* = .3016 | .0588<br>*p* = .3462 |
| Partial correlation | -.0049<br>*p* = .9589 | .0027<br>*p* = .9772 | .1268<br>*p* = .1788 | .0666<br>*p* = .4817 | .1154<br>*p* = .2215 | .0725<br>*p* = .4434 |
| **Lifetime PDD and**<br>**HC (n = 51)** |  |  |  |  |  |  |
| Pearson | -.1869<br>*p* = .1891 | -.1241<br>*p* = .3857 | -.2110<br>*p* = .1372 | .1417<br>*p* = .3213 | -.2023<br>*p* = .1545 | .0216<br>*p* = .8802 |
| Kendall | -.1576<br>*p* = .1072 | -.0615<br>*p* = .5262 | -.1341<br>*p* = .1671 | .0649<br>*p* = .5050 | -.1111<br>*p* = .2519 | -.0621<br>*p* = .5210 |
| Partial correlation | -.1973<br>*p* = .1837 | -.0580<br>*p* = .6984 | -.2654<sup>†</sup><br>*p* = .0714 | .1032<br>*p* = .4899 | -.2491<sup>†</sup><br>*p* = .0914 | -.0751<br>*p* = .6158 |
| **12-month PDD and**<br>**HC (n = 29)** |  |  |  |  |  |  |
| Pearson | -.2725<br>*p* = .1527 | -.2186<br>*p* = .2547 | -.1908<br>*p* = .3216 | .2566<br>*p* = .1790 | -.2401<br>*p* = .2097 | .0100<br>*p* = .9591 |
| Kendall | -.1147<br>*p* = .3873 | -.1112<br>*p* = .3983 | -.1208<br>*p* = .3579 | .0919<br>*p* = .4871 | -.1706<br>*p* = .1953 | -.1504<br>*p* = .2524 |
| Partial correlation | -.2945<br>*p* = .1529 | -.0729<br>*p* = .7290 | -.2113<br>*p* = .3107 | .3044<br>*p* = .1390 | -.2452<br>*p* = .2375 | -.0449<br>*p* = .8313 |
| **Lifetime MDD and HC (*n* = 73)** |  |  |  |  |  |  |
| Pearson | .0201<br>*p* = .8660 | -.0302<br>*p* = .7999 | .0663<br>*p* = .5773 | .0874<br>*p* = .4619 | .0560<br>*p* = .6382 | .1984<sup>†</sup><br>*p* = .0924 |
| Kendall | -.0100<br>*p* = .9014 | -.0153<br>*p* = .8489 | -.0011<br>*p* = .9886 | .0392<br>*p* = .6269 | .0145<br>*p* = .8564 | .1399<sup>†</sup><br>*p* = .0805 |
| Partial correlation | .0523<br>*p* = .6695 | .0124<br>*p* = .9197 | .0981<br>*p* = .4224 | .0838<br>*p* = .4934 | .0894<br>*p* = .4651 | .1414<br>*p* = .2466 |
| **12-month MDD and HC (*n* = 24)** |  |  |  |  |  |  |
| Pearson | -.1686<br>*p* = .4308 | -.2456<br>*p* = .2473 | -.0572<br>*p* = .7906 | .0493<br>*p* = .8191 | -.0832<br>*p* = .6991 | .0581<br>*p* = .7873 |
| Kendall | -.1168<br>*p* = .4267 | -.1416<br>*p* = .3332 | -.0690<br>*p* = .6373 | .0073<br>*p* = .9604 | -.0942<br>*p* = .5392 | .0327<br>*p* = .8233 |
| Partial correlation | -.2465<br>*p* = .2948 | -.2519<br>*p* = .2840 | -.1651<br>*p* = .4867 | -.0295<br>*p* = .9017 | -.1736<br>*p* = .4642 | .0019<br>*p* = .9937 |

*Note.* Pearson and Kendall tests were calculated depending on normality and scale of measurement. All correlations were calculated two-tailed. <sup>†</sup> *p* < .1, two-tailed (trend).

**Supplementary Table 12.** Associations Between Cortisone T2 and Cell Deformability Over the Entire Sample and According to Different Diagnostic Groups

|  | Erythrocytes | Monocytes | Neutrophils | Lymphocytes | Granulo-monocytes | Thrombocytes |
| --- | --- | --- | --- | --- | --- | --- |
| **Entire Sample**<br>**(n = 118)** |  |  |  |  |  |  |
| Pearson | -.0572<br>*p* = .5381 | .0371<br>*p* = .6897 | .1374<br>*p* = .1379 | .1071<br>*p* = .2481 | .1224<br>*p* = .1866 | -.0154<br>*p* = .8684 |
| Kendall | -.0689<br>*p* = .2730 | .0264<br>*p* = .6720 | .0737<br>*p* = .2382 | .0376<br>*p* = .5498 | .0784<br>*p* = .2099 | -.0331<br>*p* = .5959 |
| Partial correlation | .0044<br>*p* = .9626 | .0371<br>*p* = .6953 | .1202<br>*p* = .2028 | .0214<br>*p* = .8215 | .1005<br>*p* = .2874 | -.0394<br>*p* = .6769 |
| **Lifetime PDD and**<br>**HC (n = 51)** |  |  |  |  |  |  |
| Pearson | -.2004<br>*p* = .1586 | -.0273<br>*p* = .8894 | -.0090<br>*p* = .9500 | .1315<br>*p* = .3575 | -.0023<br>*p* = .9873 | -.0742<br>*p* = .6050 |
| Kendall | -.1631<sup>†</sup><br>*p* = .0953 | .0071<br>*p* = .9417 | .0150<br>*p* = .8773 | .0245<br>*p* = .8010 | .0457<br>*p* = .6374 | -.0754<br>*p* = .4355 |
| Partial correlation | -.2263<br>*p* = .1261 | .0091<br>*p* = .9516 | -.0073<br>*p* = .9612 | .0890<br>*p* = .5520 | -.0005<br>*p* = .9873 | -.1185<br>*p* = .4275 |
| **12-month PDD and**<br>**HC (n = 29)** |  |  |  |  |  |  |
| Pearson | -.2664<br>*p* = .1624 | -.0204<br>*p* = .9162 | .1220<br>*p* = .5285 | .3803*<br>*p* = .0419 | .1059<br>*p* = .5847 | -.0278<br>*p* = .8860 |
| Kendall | -.1696<br>*p* = .2012 | .0074<br>*p* = .9551 | .1208<br>*p* = .3579 | .1665<br>*p* = .2082 | .1162<br>*p* = .3777 | -.0567<br>*p* = .6661 |
| Partial correlation | -.2828<br>*p* = .1707 | .0359<br>*p* = .8646 | .1991<br>*p* = .3400 | .3919<sup>†</sup><br>*p* = .0527 | .1835<br>*p* = .3800 | -.0334<br>*p* = .8740 |
| **Lifetime MDD and HC (n = 73)** |  |  |  |  |  |  |
| Pearson | .0441<br>*p* = .7112 | -.0285<br>*p* = .8111 | .0550<br>*p* = .6440 | .0244<br>*p* = .8375 | .0365<br>*p* = .7590 | .1051<br>*p* = .3761 |
| Kendall | -.0042<br>*p* = .9582 | -.0072<br>*p* = .9279 | .0092<br>*p* = .9090 | -.0088<br>*p* = .9127 | .0218<br>*p* = .786 | .0915<br>*p* = .2530 |
| Partial correlation | .0767<br>*p* = .5311 | .0002<br>*p* = .9984 | .0826<br>*p* = .4998 | .0075<br>*p* = .9512 | .0634<br>*p* = .6046 | .0713<br>*p* = .5602 |
| **12-month MDD and HC (n = 24)** |  |  |  |  |  |  |
| Pearson | -.1041<br>*p* = .6284 | .0013<br>*p* = .9951 | .1700<br>*p* = .4270 | .0089<br>*p* = .9672 | .1726<br>*p* = .4200 | .1473<br>*p* = .4922 |
| Kendall | -.0657<br>*p* = .6548 | .0327<br>*p* = .8233 | .1198<br>*p* = .4129 | -.0949<br>*p* = .5185 | .1087<br>*p* = .4761 | .0762<br>*p* = .6023 |
| Partial correlation | -.0927<br>*p* = .6976 | -.0050<br>*p* = .9834 | .1284<br>*p* = .5895 | -.0858<br>*p* = .7190 | .1375<br>*p* = .5631 | .0781<br>*p* = .7434 |

*Note.* Pearson and Kendall tests were calculated depending on normality and scale of measurement. All correlations were calculated two-tailed. \* $p < .05$ . <sup>†</sup> $p < .1$ , two-tailed (trend).

**Supplementary Table 13.** Mean Cell Deformation Comparisons Between High and Minimal Depressive Disorder Risk Group Assessed by PHQ-9 Scores (T1)

|  | **Erythrocytes** | **Monocytes** | **Neutrophils** | **Lymphocytes** | **Granulo-Monocytes** | **Thrombocytes** |
| --- | --- | --- | --- | --- | --- | --- |
| **High risk T1 (n = 26)** |  |  |  |  |  |  |
| *M* | .3365 | .0585 | .0633 | .0294 | .0634 | .0914 |
| *SD* | .0172 | .0041 | .0032 | .0017 | .0030 | .0252 |
| **Minimal risk T1 (n = 69)** |  |  |  |  |  |  |
| *M* | .3375 | .0549 | .0624 | .0287 | .0625 | .0879 |
| *SD* | .0146 | .0069 | .0043 | .0018 | .0043 | .0356 |
| *t*-value/*U*-value | *t*(93) = 0.2821 | *t*(75.238) = -3.0642** | *t*(93) = -.882 | *U* = 677.5* | *t*(64.406) = -1.095 | *U* = 707.5 |
| *p*-value | .6107 | .0015 | .1901 | .0337 | .1388 | .0573 |
| **Consistent High risk (n = 13)** |  |  |  |  |  |  |
| *M* | .3407 | .0589 | .0637 | .0292 | .0638 | .0939 |
| *SD* | .0092 | .0035 | .0030 | .0016 | .0028 | .0304 |
| **Consistent minimal risk (n = 53)** |  |  |  |  |  |  |
| *M* | .3379 | .0550 | .0628 | .0289 | .0629 | .0907 |
| *SD* | .0144 | .0067 | .0043 | .0015 | .0042 | .0395 |
| *t*-value/*U*-value | *t*(64) = -.6634 | *t*(37.138) = -2.9601** | *t*(64) = -.6509 | *U* = 289 | *t*(64) = -.676 | *U* = 284 |
| *p*-value | .2547 | .0027 | .2587 | .1876 | .2507 | .1667 |

*Note.* One-tailed *t*-tests and Mann-Whitney *U* tests were calculated depending on normality and scale of measurement. \* $p < .05$ , one-tailed. \*\* $p < .01$ , one-tailed.

**Supplementary Table 14.** Associations Between Cortisol T1, T2 and T2-T1 and Cell Count Over the Entire Sample and According to Different Diagnostic Groups

|  | **Erythrocytes** | **Monocytes** | **Neutrophils** | **Lymphocytes** | **Granulo-monocytes** |
| --- | --- | --- | --- | --- | --- |
| **Entire Sample**<br>**(n = 116)** |  |  |  |  |  |
| Cortisol T1 | .0679<br>*p* = .469 | -.0196<br>*p* = .8348 | .1045<br>*p* = .2643 | .0430<br>*p* = .6464 | .1347<br>*p* = .1496 |
| Cortisol T2 | .1117<br>*p* = .2325 | -.0404<br>*p* = .6669 | .0465<br>*p* = .6199 | .0639<br>*p* = .4956 | .0494<br>*p* = .5985 |
| Cortisol T2-T1 | .0807<br>*p* = .3894 | -.0342<br>*p* = .7159 | -.0432<br>*p* = .7888 | .0419<br>*p* = .6554 | -.0697<br>*p* = .4569 |
| **Lifetime PDD and**<br>**HC (n = 50)** |  |  |  |  |  |
| Cortisol T1 | -.1574<br>*p* = .275 | .1630<br>*p* = .2579 | .2296<br>*p* = .1087 | .1925<br>*p* = .1805 | .2948<br>*p* = .03769* |
| Cortisol T2 | .0360<br>*p* = .8041 | .0746<br>*p* = .6068 | .1832<br>*p* = .2029 | .2047<br>*p* = .1539 | .2054<br>*p* = .1525 |
| Cortisol T2-T1 | .2464<br>*p* = .0846 | -.0971<br>*p* = .5022 | -.0263<br>*p* = .8562 | .0502<br>*p* = .7291 | -.0759<br>*p* = .6003 |
| **12-month PDD and**<br>**HC (n = 29)** |  |  |  |  |  |
| Cortisol T1 | -.0329<br>*p* = .8655 | .1537<br>*p* = .4261 | .4587<br>*p* = .0123* | .0885<br>*p* = .6482 | .5240<br>*p* = .0035**<sup>a</sup> |
| Cortisol T2 | -.0256<br>*p* = .8953 | .0526<br>*p* = .7865 | .3650<br>*p* = .0515<sup>†</sup> | .2068<br>*p* = .2818 | .3759<br>*p* = .0445* |
| Cortisol T2-T1 | .0032<br>*p* = .9867 | -.1107<br>*p* = .5676 | -.0330<br>*p* = .865 | .1884<br>*p* = .3276 | -.0965<br>*p* = .6185 |
| **Lifetime MDD and**<br>**HC (n = 71)** |  |  |  |  |  |
| Cortisol T1 | -.0780 | -.0106 | .1603 | .0124 | .1902 |

|  | Erythrocytes | Monocytes | Neutrophils | Lymphocytes | Granulo-monocytes |
| --- | --- | --- | --- | --- | --- |
| | $p = .5073$ | $p = .9303$ | $p = .1818$ | $p = .9181$ | $p = .112$ |
| Cortisol T2 | -.0069 | -.0926 | .1089 | .0334 | .0875 |
| | $p = .9543$ | $p = .4425$ | $p = .3659$ | $p = .7819$ | $p = .4682$ |
| Cortisol T2-T1 | .0744 | -.1141 | -.0207 | .0322 | -.0810 |
| | $p = .5375$ | $p = .3434$ | $p = .8642$ | $p = .79$ | $p = .502$ |
| **12-month MDD and HC ($n = 24$)** |  |  |  |  |  |
| Cortisol T1 | -.1567 | .2566 | .2693 | .2914 | .3236 |
| | $p = .4647$ | $p = .2262$ | $p = .2031$ | $p = .1671$ | $p = .123$ |
| Cortisol T2 | -.1640 | .0035 | .2191 | .1908 | .1868 |
| | $p = .4438$ | $p = .9872$ | $p = .3037$ | $p = .3719$ | $p = .3822$ |
| Cortisol T2-T1 | -.0594 | -.2563 | .0183 | -.0419 | -.0799 |
| | $p = .7827$ | $p = .2268$ | $p = .9323$ | $p = .8459$ | $p = .7106$ |

Note. All Pearson correlations were calculated two-tailed. <sup>a</sup> Result survived Holm-Bonferroni correction. \*\* $p < .01$ . \* $p < .05$ . <sup>†</sup> $p < .1$ , two-tailed (trend).

**Supplement Table 15.** Associations Between Cortisone T1, T2 and T2-T1 and Cell Count Over the Entire Sample and According to Different Diagnostic Groups

|  | **Erythrocytes** | **Monocytes** | **Neutrophils** | **Lymphocytes** | **Granulo-monocytes** |
| --- | --- | --- | --- | --- | --- |
| **Entire Sample**<br>**(n = 118)** |  |  |  |  |  |
| Cortisone T1 | .0500<br>*p* = .5909 | .0425<br>*p* = .6478 | .0104<br>*p* = .9109 | .0916<br>*p* = .9109 | .0559<br>*p* = .5477 |
| Cortisone T2 | .0901<br>*p* = .332 | -.0320<br>*p* = .7306 | -.0181<br>*p* = .8455 | .1071<br>*p* = .2483 | -.0123<br>*p* = .8948 |
| Cortisone T2-T1 | .0704<br>*p* = .449 | -.1008<br>*p* = .2773 | -.0399<br>*p* = .6679 | .0432<br>*p* = .6422 | -.0885<br>*p* = .3404 |
| **Lifetime PDD and**<br>**HC (n = 51)** |  |  |  |  |  |
| Cortisone T1 | -.1567<br>*p* = .2722 | .1343<br>*p* = .3473 | .1594<br>*p* = .2639 | .3036<br>*p* = .0303* | .2149<br>*p* = .13 |
| Cortisone T2 | -.0209<br>*p* = .884 | -.0140<br>*p* = .9224 | .0808<br>*p* = .5732 | .2758<br>*p* = .04996* | .0780<br>*p* = .5862 |
| Cortisone T2-T1 | .1795<br>*p* = .2075 | -.1974<br>*p* = .1649 | -.1017<br>*p* = .4778 | -.0272<br>*p* = .8498 | -.1790<br>*p* = .2089 |
| **12-month PDD and**<br>**HC (n = 29)** |  |  |  |  |  |
| Cortisone T1 | -.0119<br>*p* = .9513 | .1414<br>*p* = .4645 | .4697<br>*p* = .0102* | .1463<br>*p* = .4488 | .5307<br>*p* = .0031**<sup>a</sup> |
| Cortisone T2 | -.0025<br>*p* = .9897 | -.0548<br>*p* = .7778 | .3808<br>*p* = .0416* | .2433<br>*p* = .2034 | .3452<br>*p* = .0666<sup>†</sup> |
| Cortisone T2-T1 | .0129<br>*p* = .9469 | -.2940<br>*p* = .1217 | -.0478<br>*p* = .8054 | .1909<br>*p* = .3212 | -.1944<br>*p* = .3123 |
| **Lifetime MDD and**<br>**HC (n = 73)** |  |  |  |  |  |
| Cortisone T1 | -.1274 | .0509 | .0263 | .0369 | .0724 |

|  | Erythrocytes | Monocytes | Neutrophils | Lymphocytes | Granulo-monocytes |
| --- | --- | --- | --- | --- | --- |
| | $p = .2829$ | $p = .6691$ | $p = .8255$ | $p = .7567$ | $p = .5425$ |
| Cortisone T2 | -.0565 | -.0555 | -.0186 | .1020 | -.0237 |
| | $p = .6352$ | $p = .641$ | $p = .8759$ | $p = .3904$ | $p = .8419$ |
| Cortisone T2-T1 | .0878 | -.1609 | -.0669 | .1127 | -.1400 |
| | $p = .4602$ | $p = .1739$ | $p = .5741$ | $p = .3423$ | $p = .2375$ |
| **12-month MDD and HC ($n = 24$)** |  |  |  |  |  |
| Cortisone T1 | -.0864 | .1367 | .1807 | .2864 | .2099 |
| | $p = .6882$ | $p = .5242$ | $p = .398$ | $p = .1748$ | $p = .3248$ |
| Cortisone T2 | -.2340 | -.0486 | .1640 | .2249 | .1320 |
| | $p = .271$ | $p = .8217$ | $p = .4438$ | $p = .2906$ | $p = .5386$ |
| Cortisone T2-T1 | -.1893 | -.2457 | -.0279 | -.0897 | -.1085 |
| | $p = .3758$ | $p = .2471$ | $p = .8971$ | $p = .6767$ | $p = .6138$ |

Note. All Pearson correlations were calculated two-tailed. <sup>a</sup> Result survived Holm-Bonferroni correction. \*\* $p < .01$ . \* $p < .05$ . <sup>†</sup> $p < .1$ , two-tailed (trend).

## Supplementary Text 1

Drug intake, measured by the subject questionnaire, was analysed by forming six groups, which are: no medication (*n* = 76), psychopharmaceutical medication (*n* = 14), antihypertensive drugs (*n* = 16), thyroid dysfunction medication (*n* = 16), others (*n* = 4) and an intake combination group of the preceding categories (*n* = 10). ANOVA revealed no significant difference between the medication intake groups and cell deformation for each cell type [red blood cells (*F*(5, 130) = 1.163, *p* = .331), monocytes (*F*(5, 130) = 1.112, *p* = .357), neutrophils (*F*(5, 130) = .722, *p* = .608), lymphocytes (*F*(5, 130) = .935, *p* = .461), granulo-monocytes (*F*(5, 130) = .897, *p* = .486), thrombocytes (*F*(5, 130) = 1.027, *p* = .405)].

**Supplementary Figure 1.**
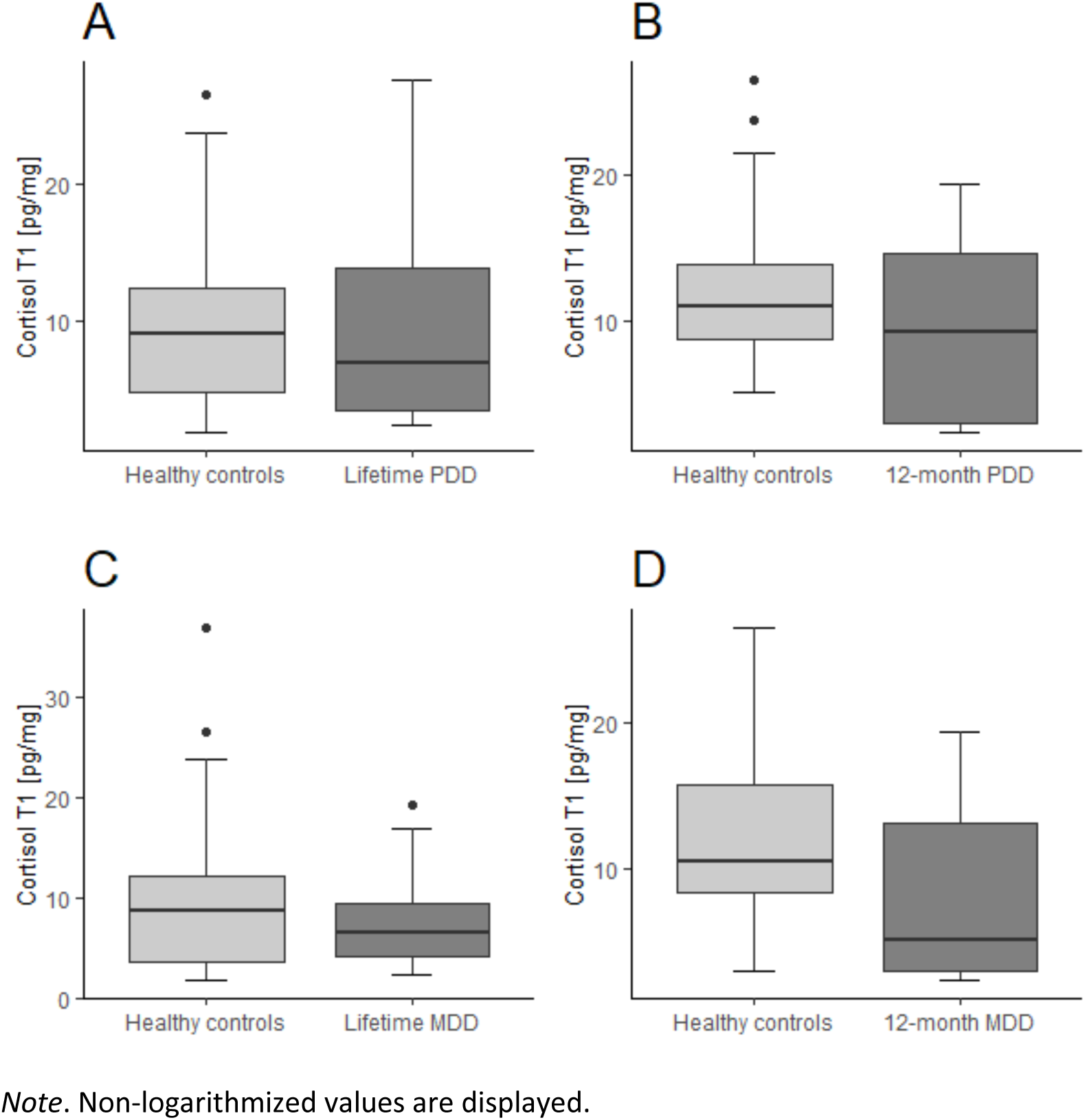
Combined Boxplots for Cortisol (T1) According to Subgroups with Differing Depressive Disorders and Healthy Controls

**Supplementary Figure 2.**
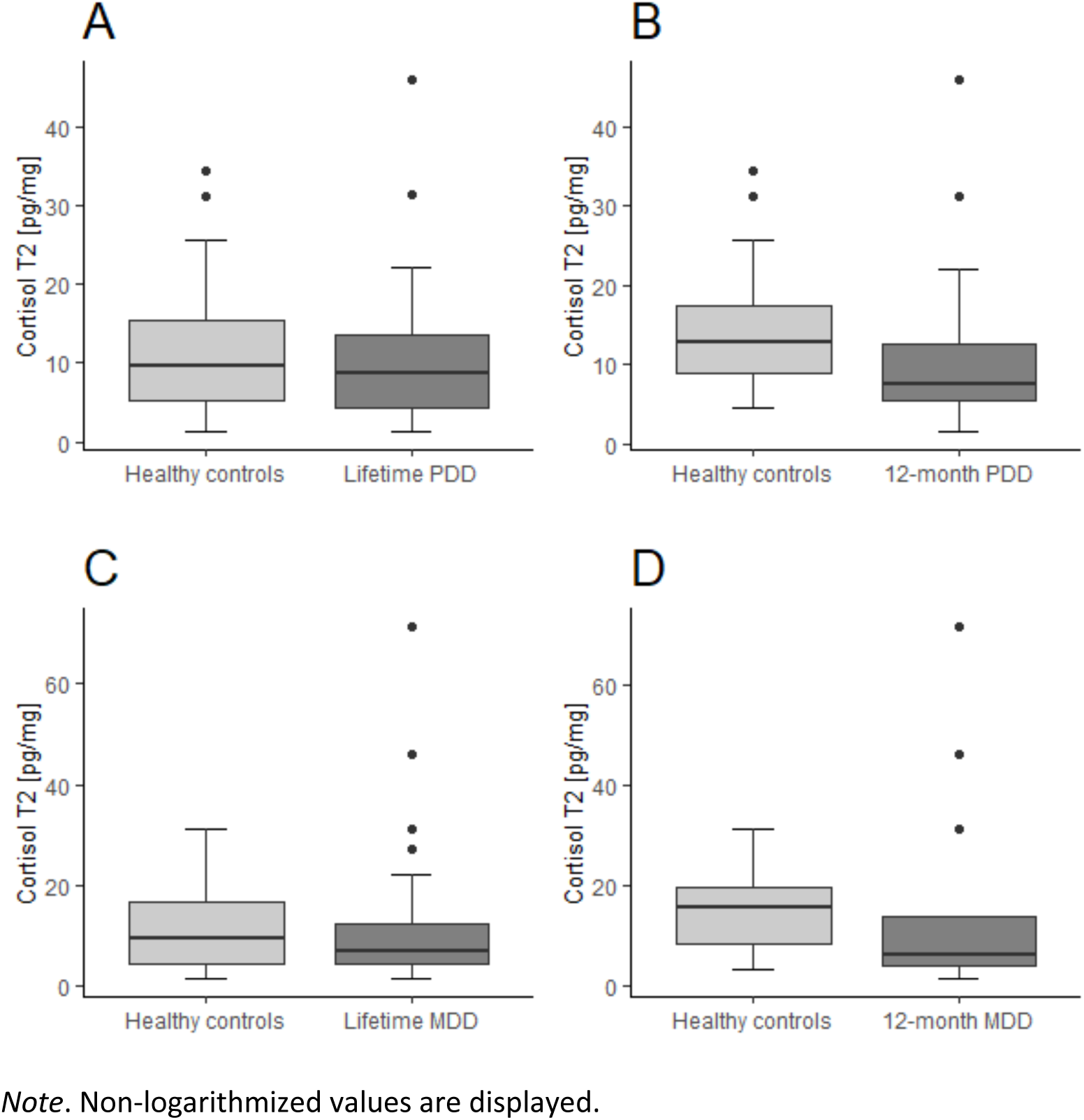
Combined Boxplots for Cortisol (T2) According to Subgroups with Differing Depressive Disorders and Healthy Controls

**Supplementary Figure 3.**
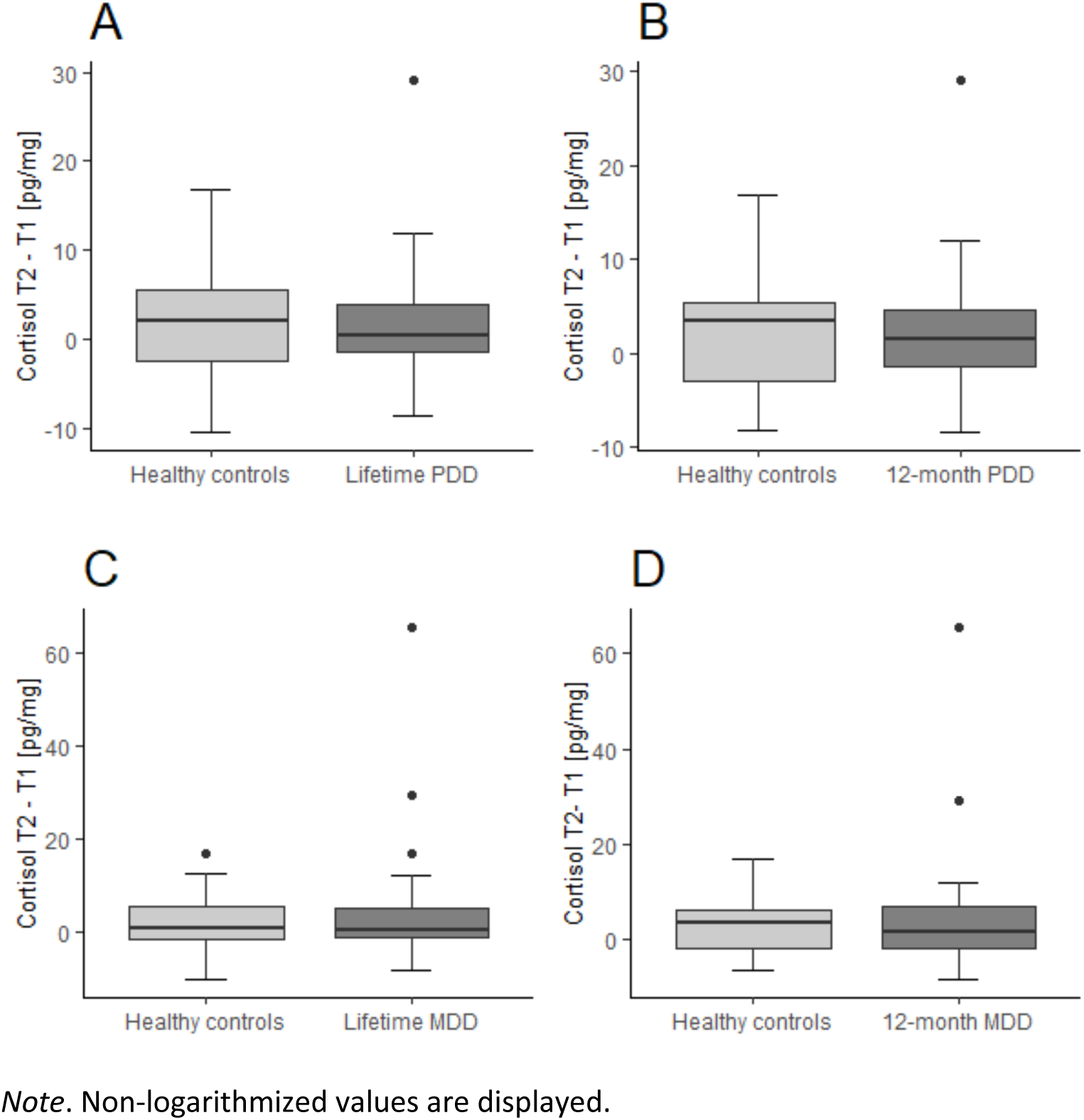
Combined Boxplots for Cortisol (T2-T1) According to Subgroups with Differing Depressive Disorders and Healthy Controls

**Supplementary Figure 4.**
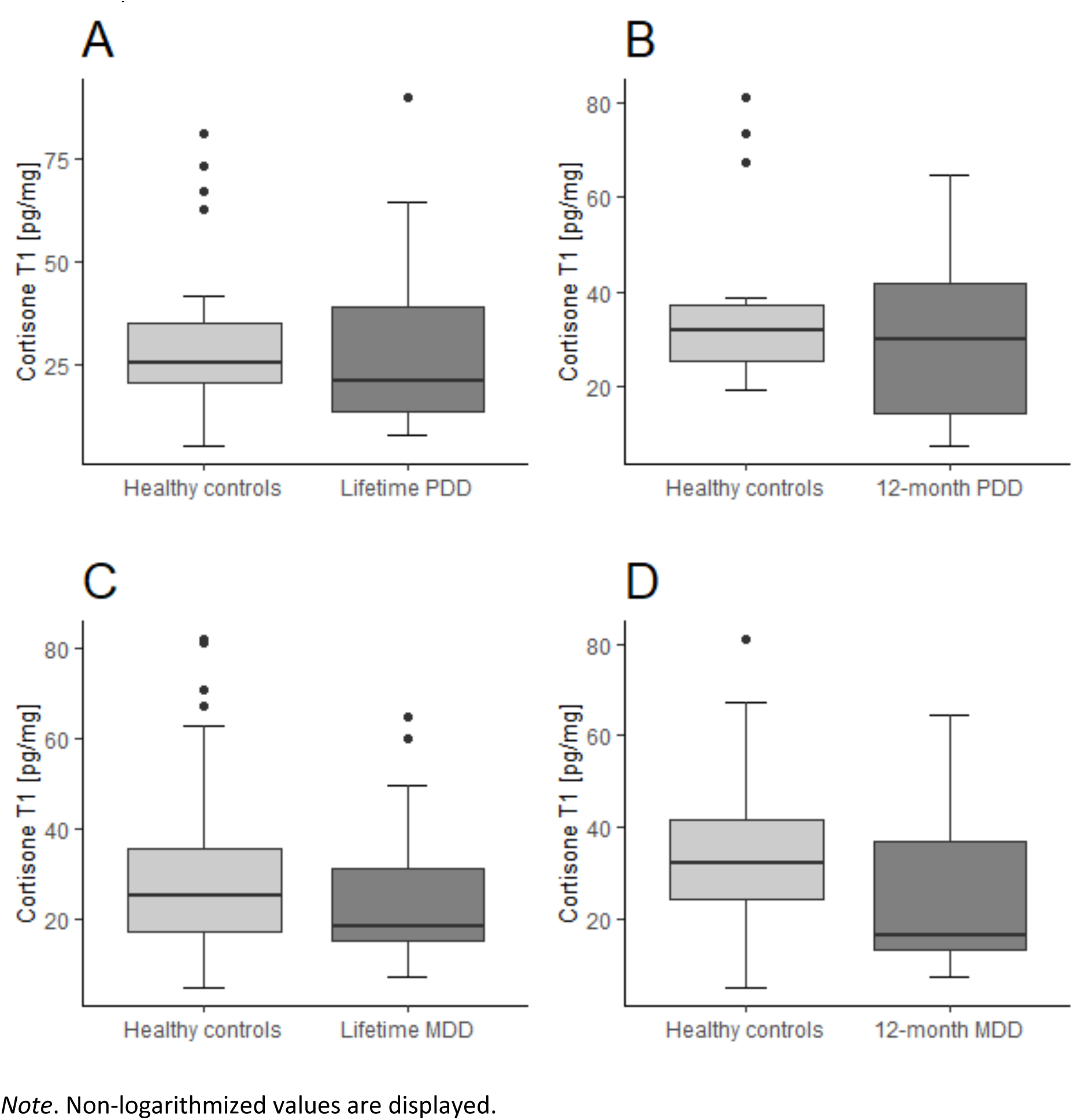
Combined Boxplots for Cortisone (T1) According to Subgroups with Differing Depressive Disorders and Healthy Controls

**Supplementary Figure 5.**
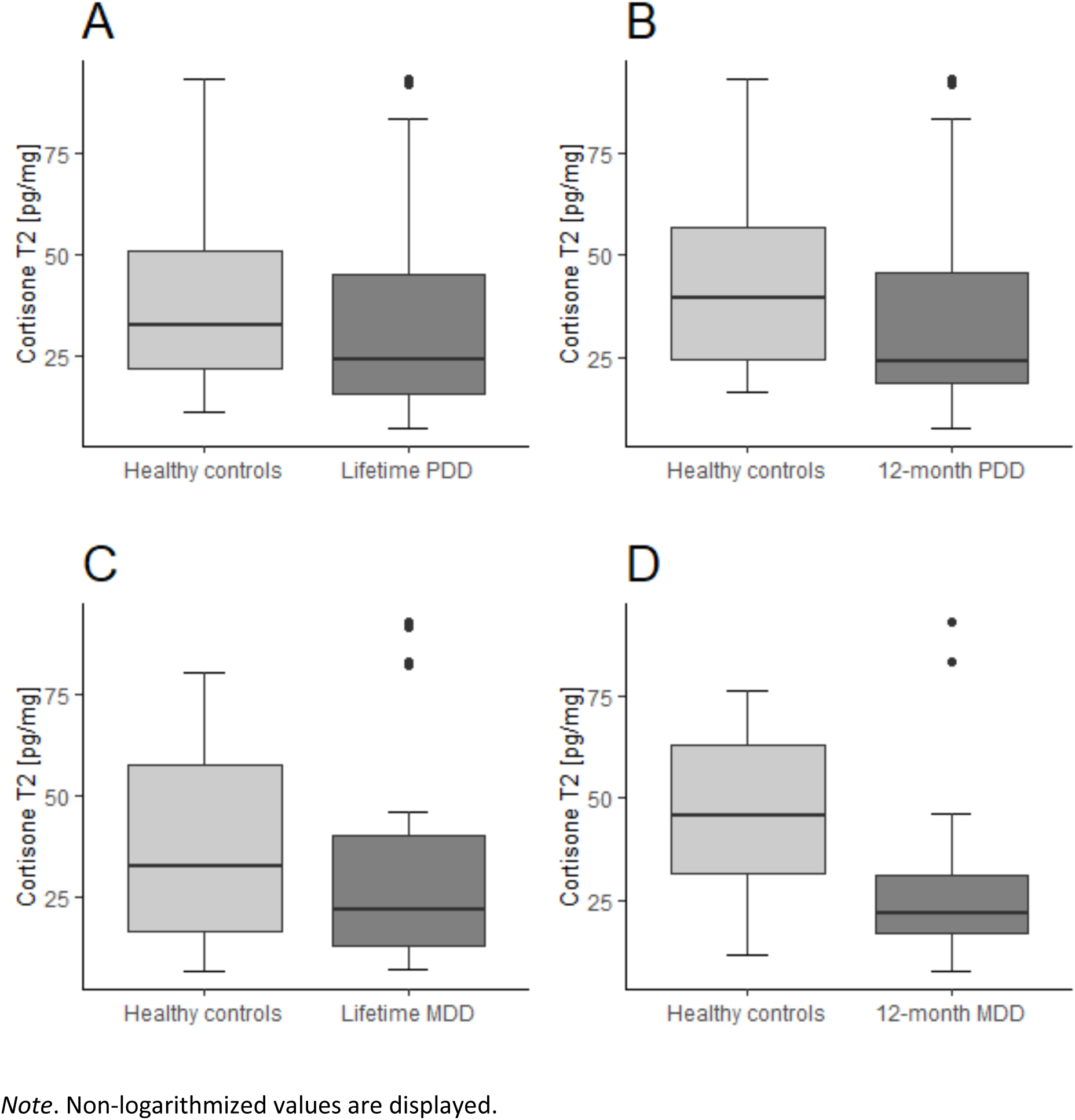
Combined Boxplots for Cortisone (T2) According to Subgroups with Differing Depressive Disorders and Healthy Controls

**Supplementary Figure 6.**
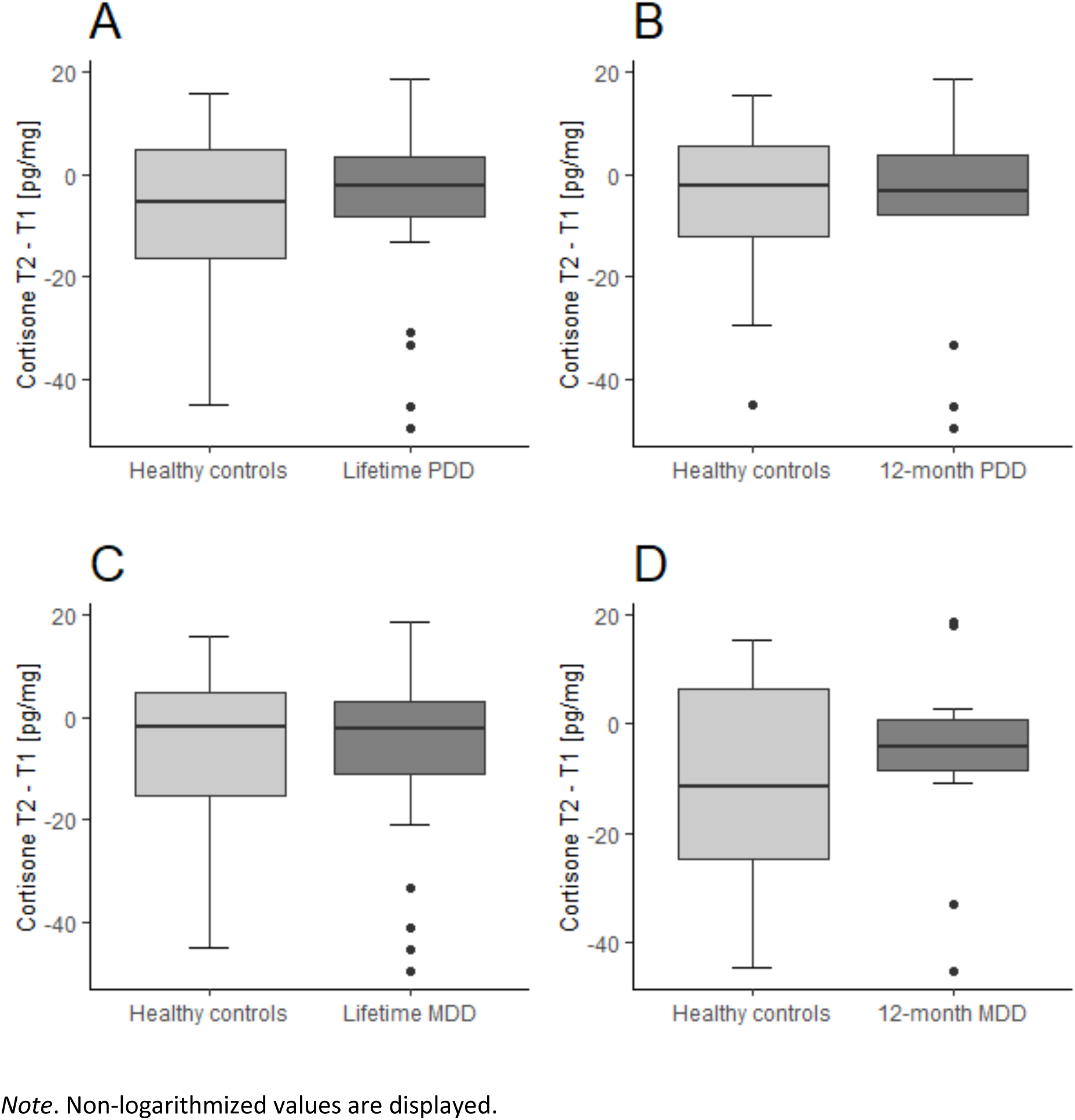
Combined Boxplots for Cortisone (T2-T1) According to Subgroups with Differing Depressive Disorders and Healthy Controls

## References

American Psychiatric Association, 2013. Diagnostic and statistical manual of mental disorders 5. Am Psychiatric Assoc 21, 591–643.

Bashant, K.R., Topefner, N., Day, C.J., Mehta, N.N., Kaplan, M.J., Summers, C., Guck, J., Chilvers, E.R., 2020. The mechanics of myeloid cells. Biol Cell 1–10. https://doi.org/10.1111/boc.201900084

Bashant, K.R., Vassallo, A., Herold, C., Berner, R., Menschner, L., Subburayalu, J., Kaplan, M.J., Summers, C., Guck, J., Chilvers, E.R., 2019. Real-time deformability cytometry reveals sequential contraction and expansion during neutrophil priming. J Leukoc Biol 105, 1143–1153.

Biselli, T., Lange, S., Sablottny, L., Steffen, J., Walther, A., 2019. Optogenetic and chemogenetic insights into the neurocircuitry of depression-like behaviour: A systematic review. European Journal of Neuroscience accepted, 327–331.

Cantave, C.Y., Ouellet-Morin, I., Giguère, C.-É., Lupien, S.J., Juster, R.-P., Geoffrion, S., Marin, M.-F., 2022. The Association of Childhood Maltreatment, Sex, and Hair Cortisol Concentrations With Trajectories of Depressive and Anxious Symptoms Among Adult Psychiatric Inpatients. Psychosom Med 84, 20–28.

Croux, C., Dehon, C., 2010. Influence functions of the Spearman and Kendall correlation measures. Stat Methods Appt 19, 497–515.

Dantzer, R., 2009. Cytokine, sickness behavior, and depression. Immunology and Allergy Clinics 29, 247–264.

Demirkan, A., Isaacs, A., Ugocsai, P., Liebisch, G., Struchalin, M., Rudan, I., Wilson, J.F., Pramstaller, P.P., Gyllensten, U., Campbell, H., Schmitz, G., Oostra, B.A., van Duijn, C.M., 2013. Plasma phosphatidylcholine and sphingomyelin concentrations are associated with depression and anxiety symptoms in a Dutch family-based lipidomics study Ays. J Psychiatr Res 47, 357–362. https://doi.org/10.1016/j.jpsychires.2012.11.001

Ekpenyong, A.E., Whyte, G., Chalut, K., Pagliara, S., Lautenschläger, F., Fiddler, C., Paschke, S., Keyser, U.F., Chilvers, E.R., Guck, J., 2012. Viscoelastic properties of differentiating blood cells are fate-and function-dependent.

Fay, M.E., Myers, D.R., Kumar, A., Turbyfield, C.T., Byler, R., Crawford, K., Mannino, R.G., Laohapant, A., Tyburski, E.A., Sakurai, Y., Rosenbluth, M.J., Switz, N.A., Sulchek, T.A., Graham, M.D., Lam, W.A., 2016. Cellular softening mediates leukocyte demargination and trafficking, thereby increasing clinical blood counts. Proc Natl Acad Sci U S A 113, 1987–1992. https://doi.org/10.1073/pnas.1508920113

Fiacco, S., Walther, A., Ehlert, U., 2019. Steroid secretion in healthy aging. Psychoneuroendocrinology 105, 64–78. https://doi.org/10.1016/j.psyneuen.2018.09.035

Gao, W., Kirschbaum, C., Grass, J., Stalder, T., 2016. LC – MS based analysis of endogenous steroid hormones in human hair. Journal of Steroid Biochemistry & Molecular Biology 162, 92–99. https://doi.org/10.1016/j.jsbmb.2015.12.022

Gao, W., Stalder, T., Foley, P., Rauh, M., Deng, H., Kirschbaum, C., 2013. Quantitative analysis of steroid hormones in human hair using a column-switching LC – APCI – MS / MS assay. Journal of Chromatography B 928, 1–8. https://doi.org/10.1016/j.jchromb.2013.03.008

Gerritsen, L., Staufenbiel, S.M., Penninx, B.W.J.H., van Hemert, A.M., Noppe, G., de Rijke, Y.B., van Rossum, E.F.C., 2019. Long-term glucocorticoid levels measured in hair in patients with depressive and anxiety disorders. Psychoneuroendocrinology 101, 246– 252.

Goecke, I.A., Alvarez, C., Henríquez, J., Salas, K., Molina, M.L., Ferreira, A., Gatica, H., 2007. Methotrexate regulates the expression of glucocorticoid receptor alpha and beta isoforms in normal human peripheral mononuclear cells and human lymphocyte cell lines in vitro. Mol Immunol 44, 2115–2123.

Goldsmith, D.R., Rapaport, M.H., Miller, B.J., 2016. A meta-analysis of blood cytokine network alterations in psychiatric patients: comparisons between schizophrenia, bipolar disorder and depression. Mol Psychiatry 1–14. https://doi.org/10.1038/mp.2016.3

Guidi, J., Fava, G.A., Fava, M., Papakostas, G.I., 2011. Efficacy of the sequential integration of psychotherapy and pharmacotherapy in major depressive disorder: A preliminary meta-analysis. Psychol Med 41, 321–331. https://doi.org/10.1017/S0033291710000826

Hasin, D.S., Sarvet, A.L., Meyers, J.L., Saha, T.D., Ruan, W.J., Stohl, M., Grant, B.F., 2018. Epidemiology of adult DSM-5 major depressive disorder and its specifiers in the United States. JAMA Psychiatry 75, 336–346. https://doi.org/10.1001/jamapsychiatry.2017.4602

He, C., Levis, B., Riehm, K.E., Saadat, N., Levis, A.W., Azar, M., Rice, D.B., Krishnan, A., Wu, Y., Sun, Y., Imran, M., Boruff, J., Cuijpers, P., Gilbody, S., Ioannidis, J.P.A., Kloda, L.A., McMillan, D., Patten, S.B., Shrier, I., Ziegelstein, R.C., Akena, D.H., Arroll, B., Ayalon, L., Baradaran, H.R., Baron, M., Beraldi, A., Bombardier, C.H., Butterworth, P., Carter, G., Chagas, M.H.N., Chan, J.C.N., Cholera, R., Clover, K., Conwell, Y., de Man-Van Ginkel, J.M., Fann, J.R., Fischer, F.H., Fung, D., Gelaye, B., Goodyear-Smith, F., Greeno, C.G., Hall, B.J., Harrison, P.A., Härter, M., Hegerl, U., Hides, L., Hobfoll, S.E., Hudson, M., Hyphantis, T.N., Inagaki, M., Ismail, K., Jetté, N., Khamseh, M.E., Kiely, K.M., Kwan, Y., Lamers, F., Liu, S.I., Lotrakul, M., Loureiro, S.R., Löwe, B., Marsh, L., McGuire, A., Mohd-Sidik, S., Munhoz, T.N., Muramatsu, K., Osório, F.L., Patel, V., Pence, B.W., Persoons, P., Picardi, A., Reuter, K., Rooney, A.G., da Silva Dos Santos, I.S., Shaaban, J., Sidebottom, A., Simning, A., Stafford, L., Sung, S., Tan, P.L.L., Turner, A., van Weert, H.C.P.M., White, J., Whooley, M.A., Winkley, K., Yamada, M., Thombs, B.D., Benedetti, A., 2020. The Accuracy of the Patient Health Questionnaire-9 Algorithm for Screening to Detect Major Depression: An Individual Participant Data Meta-Analysis. Psychother Psychosom 89, 25–37. https://doi.org/10.1159/000502294

Heiske, A., Jesberg, J., Krieg, J.-C., Vedder, H., 2003. Differential effects of antidepressants on glucocorticoid receptors in human primary blood cells and human monocytic U-937 cells. Neuropsychopharmacology 28, 807–817.

Hinkelmann, K., Muhtz, C., Dettenborn, L., Agorastos, A., Wingenfeld, K., Spitzer, C., Gao, W., Kirschbaum, C., Wiedemann, K., Otte, C., 2013. Association between childhood trauma and low hair cortisol in depressed patients and healthy control subjects. Biol Psychiatry 74, e15–e17.

Holm, S., 1979. A Simple Sequentially Rejective Multiple Test ProcedureAuthor. Scandinavian Journal of Statistics 6, 65–70.

Hoyer, J., Voss, C., Strehle, J., Venz, J., Pieper, L., Wittchen, H.-U., Ehrlich, S., Beesdo-Baum, K., 2020. Test-retest reliability of the computer-assisted DIA-X-5 interview for mental disorders. BMC Psychiatry 20, 1–16.

Iob, E., Kirschbaum, C., Steptoe, A., 2020. Persistent depressive symptoms, HPA-axis hyperactivity, and inflammation: the role of cognitive-affective and somatic symptoms. Mol Psychiatry 25, 1130–1140.

Jasenovec, T., Radosinska, D., Celusakova, H., Filcikova, D., Babinska, K., Ostatnikova, D., Radosinska, J., 2019. Erythrocyte deformability in children with autism spectrum disorder: correlation with clinical features. Physiological Res 68, S307–S313.

Kim, J., Shin, Y.J., Ha, L.J., Kim, Deok-Ho, Kim, Dong-Hwee, 2019. Unraveling the mechanobiology of the immune system. Adv Healthc Mater 8, 1801332.

Knowles, E.E.M., Huynh, K., Meikle, P.J., Göhring, H.H.H., Olvera, R.L., Mathias, S.R., Meikle, P.J., Go, H.H.H., Duggirala, R., Almasy, L., Blangero, J., Curran, J.E., Glahn, D.C., 2017. The lipidome in major depressive disorder : Shared genetic influence for ether-phosphatidylcholines, a plasma-based phenotype related to inflammation, and disease risk. European Psychiatry 43, 44–50.

Kräter, M., Abuhattum, S., Soteriou, D., Jacobi, A., Krüger, T., Guck, J., Herbig, M., 2021. AIDeveloper: deep learning image classification in life science and beyond. Advanced Science 2003743, 1–12. https://doi.org/10.1101/2020.03.03.975250

Kroenke, K., Spitzer, R.L., Williams, J.B.W., 2001. The phq-9. J Gen Intern Med 16, 606–613.

Kubánková, M., Hohberger, B., Hoffmanns, J., Fürst, J., Herrmann, M., Guck, J., Kräter, M., 2021. Physical phenotype of blood cells is altered in COVID-19. Biophys J 120, 2838– 2847.

Lam, W.A., Rosenbluth, M.J., Fletcher, D.A., 2008. Increased leukaemia cell stiffness is associated with symptoms of leucostasis in paediatric acute lymphoblastic leukaemia. Br J Haematol 142, 497.

Levis, B., Sun, Y., He, C., Wu, Y., Krishnan, A., Bhandari, P.M., Neupane, D., Imran, M., Brehaut, E., Negeri, Z., Fischer, F.H., Benedetti, A., Thombs, B.D., 2020. Accuracy of the PHQ-2 Alone and in Combination with the PHQ-9 for Screening to Detect Major Depression: Systematic Review and Meta-analysis. JAMA - Journal of the American Medical Association 323, 2290–2300. https://doi.org/10.1001/jama.2020.6504

Li, L., Leung, D.Y.M., Hall, C.F., Goleva, E., 2006. Divergent expression and function of glucocorticoid receptor β in human monocytes and T cells. J Leukoc Biol 79, 818–827.

Liu, X., Zheng, P., Zhao, X., Zhang, Y., Hu, C., Li, J., Zhao, J., Zhou, J., Xie, P., Xu, G., 2016. Plasma lipidomics reveals potential lipid markers of major depressive disorder. Anal Bioanal Chem 408, 6497–6507. https://doi.org/10.1021/acs.jproteome.5b00144

Lopez-Vilchez, I., Diaz-Ricart, M., Navarro, V., Torramade, S., Zamorano-Leon, J., Lopez-Farre, A., Galan, A.M., Gasto, C., Escolar, G., 2016. Endothelial damage in major depression patients is modulated by SSRI treatment, as demonstrated by circulating biomarkers and an in vitro cell model. Transl Psychiatry 6, e886–e886.

Lu, K.D., Radom-Aizik, S., Haddad, F., Zaldivar, F., Kraft, M., Cooper, D.M., 2017. Glucocorticoid receptor expression on circulating leukocytes differs between healthy male and female adults. J Clin Transl Sci 1, 108–114.

Lynall, M.-E., Turner, L., Bhatti, J., Cavanagh, J., Boer, P. de, Mondelli, V., Jones, D., Drevets, W.C., Cowen, P., Harrison, N.A., Pariante, C.M., Pointon, L., Consortium, N., Clatworthy, M.R., Bullmore, E., 2019. Peripheral blood cell immunophenotyping reveals distinct subgroups of inflamed depression. Biol Psychiatry 706309. https://doi.org/10.1101/706309

Miller, R., Kirschbaum, C., 2019. Cultures under stress: A cross-national meta-analysis of cortisol responses to the Trier Social Stress Test and their association with anxiety-related value orientations and internalizing mental disorders. Psychoneuroendocrinology 105, 147–154. https://doi.org/10.1016/j.psyneuen.2018.12.236

Moylan, S., Berk, M., Dean, O.M., Samuni, Y., Williams, L.J., O’Neil, A., Hayley, A.C., Pasco, J.A., Anderson, G., Jacka, F.N., 2014. Oxidative & nitrosative stress in depression: why so much stress? Neurosci Biobehav Rev 45, 46–62.

Moylan, S., Maes, M., Wray, N.R., Berk, M., 2013. The neuroprogressive nature of major depressive disorder: pathways to disease evolution and resistance, and therapeutic implications. Mol Psychiatry 18, 595–606.

Müller, C.P., Reichel, M., Mühle, C., Rhein, C., Gulbins, E., Kornhuber, J., 2015. Membrane lipids in major depression and anxiety disorders ☆. Biochimica et Biophysica Acta Brain 1851, 1052–1065. https://doi.org/10.1016/j.bbalip.2014.12.014

Munder, T., Flückiger, C., Leichsenring, F., Abbass, A.A., Hilsenroth, M.J., Luyten, P., Rabung, S., Steinert, C., Wampold, B.E., 2019. Is psychotherapy effective? A re-analysis of treatments for depression. Epidemiol Psychiatr Sci 28, 268–274.

Munkholm, K., Paludan-Müller, A.S., Boesen, K., 2019. Considering the methodological limitations in the evidence base of antidepressants for depression: a reanalysis of a network meta-analysis. BMJ Open 9, e024886.

Nawaz, A.A., Urbanska, M., Herbig, M., Nötzel, M., Kräter, M., Rosendahl, P., Herold, C., Toepfner, N., Kubánková, M., Goswami, R., Abuhattum, S., Reichel, F., Müller, P., Taubenberger, A., Girardo, S., Jacobi, A., Guck, J., 2020. Intelligent image-based deformation-assisted cell sorting with molecular specificity. Nat Methods 17, 595–599. https://doi.org/10.1038/s41592-020-0831-y

Otte, C., Gold, S.M., Penninx, B.W., Pariante, C.M., Etkin, A., Fava, M., Mohr, D.C., Schatzberg, A.F., 2016. Major depressive disorder. Nature Publishing Group 2, 1–21. https://doi.org/10.1038/nrdp.2016.65

Otto, O., Rosendahl, P., Mietke, A., Golfier, S., Herold, C., Klaue, D., Girardo, S., Pagliara, S., Ekpenyong, A., Jacobi, A., Wobus, M., Töpfner, N., Keyser, U.F., Mansfeld, J., Fischer-Friedrich, E., Guck, J., 2015. Real-time deformability cytometry: on-the-fly cell mechanical phenotyping. Nat Methods 12, 199.

Pariante, C.M., 2017. Why are depressed patients inflamed? A reflection on 20 years of research on depression, glucocorticoid resistance and inflammation. European Neuropsychopharmacology 27, 554–559. https://doi.org/10.1016/j.euroneuro.2017.04.001

Penz, M., Kanthak, M.K., Pieper, L., Beesdo-baum, K., Walther, A., Miller, R., Stalder, T., Kirschbaum, C., 2018. The Dresden Burnout Study: Study protocol of a prospective cohort study for the bio-psychological investigation of burnout Marlene Penz. Int J Methods Psychiatr Res 1–11. https://doi.org/10.1002/mpr.1613

Psarraki, E.E., Kokka, I., Bacopoulou, F., Chrousos, G.P., Artemiadis, A., Darviri, C., 2021. Is there a relation between major depression and hair cortisol? A systematic review and meta-analysis. Psychoneuroendocrinology 124, 105098.

Ravetto, A., Wyss, H.M., Anderson, P.D., den Toonder, J.M.J., Bouten, C.V.C., 2014. Monocytic cells become less compressible but more deformable upon activation. PLoS One 9, e92814.

Rodrigues, R., Petersen, R.B., Perry, G., 2014. Parallels between major depressive disorder and Alzheimer’s disease: role of oxidative stress and genetic vulnerability. Cell Mol Neurobiol 34, 925–949.

Ronchetti, S., Ricci, E., Migliorati, G., Gentili, M., Riccardi, C., 2018. How glucocorticoids affect the neutrophil life. Int J Mol Sci 19, 4090.

Rosenbluth, M.J., Lam, W.A., Fletcher, D.A., 2008. Analyzing cell mechanics in hematologic diseases with microfluidic biophysical flow cytometry. Lab Chip 8, 1062–1070.

Rothe, N., Ste, J., Penz, M., Kirschbaum, C., Walther, A., 2020. Examination of peripheral basal and reactive cortisol levels in major depressive disorder and the burnout syndrome : A systematic review. Neurosci Biobehav Rev 113, 1–39. https://doi.org/10.1016/j.neubiorev.2020.02.024

Rothe, N., Vogel, S., Schmelzer, K., Kirschbaum, C., Penz, M., Wekenborg, M.K., Gao, W., Walther, A., 2021. The moderating effect of cortisol and dehydroepiandrosterone on the relation between sleep and depression or burnout. Compr Psychoneuroendocrinol 7, 100051. https://doi.org/10.1016/j.cpnec.2021.100051

Rush, A.J., Trivedi, M.H., Wisniewski, S.R., Ph, D., Nierenberg, A.A., Stewart, J.W., Warden, D., Ph, D., Niederehe, G., Ph, D., Thase, M.E., Lavori, P.W., Ph, D., Lebowitz, B.D., Ph, D., Mcgrath, P.J., Rosenbaum, J.F., Sackeim, H.A., Ph, D., Kupfer, D.J., Luther, J., Fava, M., 2006. Acute and Longer-Term Outcomes in Depressed Outpatients Requiring One or Several Treatment Steps: a STAR* D report. American Journal of Psychiatry 163, 1905– 1917.

Saha, A.K., Schmidt, B.R., Wilhelmy, J., Nguyen, V., Abugherir, A., Do, J.K., Nemat-Gorgani, M., Davis, R.W., Ramasubramanian, A.K., 2019. Red blood cell deformability is diminished in patients with Chronic Fatigue Syndrome. Clin Hemorheol Microcirc 71, 113–116.

Stalder, T., Steudte-schmiedgen, S., Alexander, N., Klucken, T., Vater, A., Wichmann, S., Kirschbaum, C., Miller, R., 2017. Stress-related and basic determinants of hair cortisol in humans: A meta-analysis 77, 261–274.

Stetler, C., Miller, G., 2011. Depression and hypothalamic-pituitary-adrenal activation: A quantitative summary of four decades of research. Psychosom Med 126, 114–126. https://doi.org/10.1097/PSY.0b013e31820ad12b

Steudte-Schmiedgen, S., Kirschbaum, C., Alexander, N., Stalder, T., 2016. An integrative model linking traumatization, cortisol dysregulation and posttraumatic stress disorder: Insight from recent hair cortisol findings. Neurosci Biobehav Rev 69, 124–135.

Steudte-schmiedgen, S., Wichmann, S., Stalder, T., Hilbert, K., Muehlhan, M., Lueken, U., Beesdo-baum, K., 2017. Hair cortisol concentrations and cortisol stress reactivity in generalized anxiety disorder, major depression and their comorbidity. J Psychiatr Res 84, 184–190. https://doi.org/10.1016/j.jpsychires.2016.09.024

Team, R.C., 2017. R Core Team (2017). R: A language and environment for statistical computing. R Found. Stat. Comput. Vienna, Austria. URL http://www.R-project.org/., page R Foundation for Statistical Computing.

Toepfner, N., Herold, C., Otto, O., Rosendahl, P., Jacobi, A., Kräter, M., Stächele, J., Menschner, L., Herbig, M., Ciuffreda, L., 2018. Detection of human disease conditions by single-cell morpho-rheological phenotyping of blood. Elife 7, e29213.

Urbanska, M., Muñoz, H.E., Shaw Bagnall, J., Otto, O., Manalis, S.R., di Carlo, D., Guck, J., 2020. A comparison of microfluidic methods for high-throughput cell deformability measurements. Nat Methods 17. https://doi.org/10.1038/s41592-020-0818-8

Vos, T., Abajobir, A.A., Abbafati, C., Abbas, K.M., Abate, K.H., Abd-Allah, F., …, Murray, C.J.L., 2017. Global, regional, and national incidence, prevalence, and years lived with disability for 328 diseases and injuries for 195 countries, 1990 – 2016: a systematic analysis for the Global Burden of Disease Study 2016. Lancet 390, 1211–59. https://doi.org/10.1016/S0140-6736(17)32154-2

Walther, A., Cannistraci, C.V., Simons, K., Durán, C., Gerl, M.J., Wehrli, S., Kirschbaum, C., 2018. Lipidomics in Major Depressive Disorder. Front Psychiatry 9. https://doi.org/10.3389/fpsyt.2018.00459

Walther, A., Kirschbaum, C., Wehrli, S., Rothe, N., Wekenborg, M., Penz, M., Gao, W., 2022a. Depressive symptoms are negatively associated with hair anandamide (AEA) levels: A cross-lagged panel analysis of four annual assessment waves examining hair endocannabinoids and cortisol. Prog Neuropsychopharmacol Biol Psychiatry.

Walther, A., Mackens-Kiani, A., Eder, J., Herbig, M., Herold, C., Kirschbaum, C., Guck, J., Wittwer, L.D., Beesdo-Baum, K., Kräter, M., 2022b. Depressive disorders are associated with increased peripheral blood cell deformability: a cross-sectional case-control study (Mood-Morph). Transl Psychiatry 12, 1–12.

Ware Jr, J.E., Kosinski, M., Keller, S.D., 1996. A 12-Item Short-Form Health Survey: construction of scales and preliminary tests of reliability and validity. Med Care 220– 233.

Wennig, R., 2000. Potential problems with the interpretation of hair analysis results. Forensic Sci Int 107, 5–12.

Wilson, C., González-Billault, C., 2015. Regulation of cytoskeletal dynamics by redox signaling and oxidative stress: implications for neuronal development and trafficking. Front Cell Neurosci 9, 381.

Wolkowitz, O.M., Epel, E.S., Reus, V.I., Mellon, S.H., 2010. Depression gets old fast: do stress and depression accelerate cell aging? Depress Anxiety 27, 327–338.

Wong, G.T.-H., Chang, R.C.-C., Law, A.C.-K., 2013. A breach in the scaffold: the possible role of cytoskeleton dysfunction in the pathogenesis of major depression. Ageing Res Rev 12, 67–75.

World Health Organisation (WHO), 2017. Depression and Other Common Mental Disorders Global Health Estimates. https://doi.org/ (WHO reference number: WHO/MSD/MER/2017.2)

Wu, P.-H., Aroush, D.R.-B., Asnacios, A., Chen, W.-C., Dokukin, M.E., Doss, B.L., Durand-Smet, P., Ekpenyong, A., Guck, J., Guz, N. v, 2018. A comparison of methods to assess cell mechanical properties. Nat Methods 15, 491–498.

Zorn, J. v, Schür, R.R., Boks, M.P., Kahn, R.S., Joëls, M., Vinkers, C.H., 2017. Cortisol stress reactivity across psychiatric disorders: A systematic review and meta-analysis. Psychoneuroendocrinology 77, 25–36.

